# Cross-System Meta-Analysis of Machine Learning Predictors Identifies Value-Specific Risk Drivers and Interactions Underlying Acute Kidney Injury

**DOI:** 10.64898/2026.08.31.26361849

**Authors:** Ho Yin Chan, Deyi Li, Alan S.L. Yu, John A. Kellum, Dana Y. Fuhrman, Qi Xu, Elizabeth A. Chrischilles, Lindsay G. Cowell, Sravani Chandaka, A. Jerrod Anzalone, Jacob Kean, Kathleen M. McTigue, Abu Saleh Mohammad Mosa, Bradley Taylor, Mahanaz Syed, Lemuel R. Waitman, Yong Hu, Mei Liu

## Abstract

**Background:** Current understanding of acute kidney injury (AKI) risk factors remains largely descriptive, offering limited precision into how specific biomarker values or physiologic thresholds influence susceptibility. We aimed to synthesize knowledge from machine learning models trained across multiple health systems to identify generalizable, value-specific risk drivers and biomarker interactions contributing to AKI risk.

**Methods:** We analyzed electronic health records (EHRs) from 785,497 adult inpatients between 2010 and 2019 across nine U.S. academic medical centers within PCORnet. Interpretable gradient boosting machine models were independently developed at each health system to quantify predictor-outcome associations. Meta-regression was applied to integrate these site-level results, characterize nonlinear value-risk relationships, and identify bivariate interactions between predictors.

**Result:** Meta-analysis revealed consistent, value-specific risk drivers across health systems. An increase in glucose from 100 mg/dL to 140 mg/dL was associated with a 1.46-fold higher risk of AKI. Chloride and anion gap also demonstrated elevated AKI risk with risk increases overlapping portions of their reference ranges, with anion gap showing a 1.14-fold increase across 4–12 mmol/L and chloride a 1.28-fold increase across 96–100 mEq/L. Electrolytes including potassium, calcium, and sodium showed quadratic associations with AKI risk. Bivariate meta-regression identified interactions between key predictors, highlighting pathways that jointly modulate AKI risk.

**Conclusion:** This cross-system meta-analysis synthesizes machine learning-derived evidence into clinically interpretable knowledge, revealing how specific biomarker ranges and interactions modulate AKI risk. By moving beyond surface-level associations to quantitative, generalizable physiologic thresholds, these findings provide actionable insights to enhance risk stratification and personalized prevention in hospital care.

**Highlights:**

- Cross-system meta-analysis uncovered generalizable, value-specific AKI risk drivers
- Glucose, chloride, and anion gap within reference ranges linked to higher AKI risk
- Key predictor interactions suggest coordinated pathways jointly modulating AKI risk

## 1. Introduction

Acute kidney injury (AKI) is a life-threatening syndrome affecting 10–15% of hospitalized patients and over 50% of critically ill patients, conferring a seven-fold higher in-hospital mortality than those without AKI.[1–3] Its incidence remains high worldwide, regardless of healthcare resources, underscoring AKI as a global health challenge.[4] Survivors face elevated risks of cardiovascular disease (CVD), chronic kidney disease (CKD), and progression to end-stage renal disease (ESRD).[5] With few effective therapies available, prevention remains the most crucial strategy to reduce its burden.[6]

Accurate identification of high-risk patients is central to prevention and has motivated the development of numerous AKI prediction models.[1, 7–11] However, these models often have limited generalizability because the distribution of clinical variables varies across populations and health systems.[1] In addition, inconsistent definitions, data structures, and extraction methods hinder large-scale meta-analyses and limit reproducibility across studies.[2, 12] Importantly, most prior studies provide only surface-level associations, without specifying the biomarker ranges or interactions that drive AKI risk.[6]

To address these limitations, we integrated electronic health records (EHRs) from nine academic medical centers through a unified data extraction and modeling pipeline. The dataset includes 785,497 adults with 1,354,516 inpatient encounters from health systems across Kansas, Missouri, Nebraska, Iowa, Wisconsin, Utah, Texas, and Pennsylvania. The EHR data were standardized using the PCORnet common data model (CDM) and obtained from two clinical research networks within PCORnet: the Greater Plains Collaborative (GPC) and PaTH.[13, 14]

Leveraging this standardized dataset, we trained interpretable machine learning (ML) models at each site and then applied meta-regression^[15]^, treating site-specific models as independent studies. This framework incorporates univariate and multivariate meta-regression with random effects and causal inference to elucidate underlying mechanisms. Unlike traditional meta-analyses, this approach quantifies how specific biomarker ranges influence AKI risk.

To our knowledge, this study represents the first large-scale, multi-center profiling of AKI risk predictors using a standardized ML-meta-regression framework. By synthesizing knowledge distilled from site-specific ML models across health systems, our approach captures nonlinear associations, value-specific risk drivers, and predictor interactions. This framework moves beyond surface-level associations to provide mechanistic, generalizable insights into AKI pathophysiology, offering a scalable foundation for precision risk stratification and prevention in clinical care.

## 2. Materials and methods

### 2.1 Data Source

We extracted EHR data from nine academic medical centers within the GPC and PaTH clinical research networks, both part of PCORnet. These centers include University of Missouri Health Care (UMHC), Medical College of Wisconsin (MCW), University of Iowa Healthcare (UIOWA), University of Kansas Medical Center (KUMC), University of Nebraska Medical Center (UNMC), University of Texas Health Science Center at San Antonio (UTHSCSA), University of Texas Southwestern Medical Center (UTSW), University of Utah (UofU) and University of Pittsburgh (UPITT). The data collected were formatted in the PCORnet CDM, which provides a common representation through terminologies, vocabularies, and coding schemes. All data were de-identified according to the Health Insurance Portability and Accountability Act of 1996 (HIPAA) ‘Safe Harbor’ criteria[16]. This study was determined by the institutional review boards of the University of Florida, University of Pittsburgh (coordinating center for PaTH), and University of Missouri (coordinating center for GPC) as non-human subject research because it only involved the collection of existing and de-identified patient medical data.

### 2.2 Study Cohort and Data Processing

The study cohort included adult patients (18 ≤ age ≤ 90) with inpatient encounters (length of stay ≥ 2 days) from the years 2010 to 2019 that had at least two serum creatinine (SCr) records available during the hospital stay.[17] Patients showing evidence of severe kidney dysfunction either at or before admission were excluded based on the following criteria (a) estimated glomerular filtration rate (eGFR) < 15 mL/min/1.73 m^2^, (b) having undergone any dialysis procedure or renal transplantation (RRT) prior to the visit, (c) requiring RRT within 48 hours of their first SCr measurement record, (d) having pre-existing ESRD, (e) having sustained major burn injuries. Baseline SCr levels were determined using a stepwise approach, as illustrated in **Figure S1**. AKI onset was defined according to the SCr criteria specified in the Kidney Disease: Improving Global Outcomes (KDIGO) Clinical Practice Guideline[18].

We extracted clinical variables available in the PCORnet CDM tables, including demographic details (age, gender, race, Hispanic), diagnosis codes (ICD-9 or ICD-10), procedure codes (ICD and CPT code), lab tests (LOINC code), medications (RXNORM and NDC code) and vital signs (weight, BMI, systolic blood pressure, and diastolic blood pressure). Each medical event is timestamped relative to the hospital admission date. Details of the medical codes in this study are provided in **Table S1**.

We defined the onset day for patients with AKI and the last SCr measurement day for control patients as anchored events. For vital signs and lab test results, we used the most recent values from the 24 hours prior to the anchored events. Historical diagnosis within a year before admission and medication records between 30 days before admission and 24 hours before the anchored events were collected. Multiple measurements of vitals and labs in the same day were averaged. Diagnosis codes were categorized as before and after 6 months relative to the admission date, with ICD-10 code converted to ICD-9 code and all ICD-9 codes rolled up to the 3-digit category level. LOINC codes were rolled up to group level whenever available. Medications were converted to the Anatomical Therapeutic Chemical (ATC) code and rolled up to the 4th level. Categorical predictors were transformed into one-hot vectors.

Predictors were excluded if they were excessively missing (>95% missingness for continuous variables) or too sparse (<5% positive class prevalence for binary variables). Outliers were removed using the interquartile range (IQR) method, defined as values below *Q*1 − 1.5 × *IQR* or above *Q*3 + 1.5 × *IQR*, where *Q*1 and *Q*3 represent the 25th and 75th percentiles, respectively. The number of predictors used is listed in **Table S2**.

### 2.3 Modeling

We reserved 20% of data from each site as a stratified test set for model evaluation and trained a CatBoost model[19], a gradient boosting machine algorithm, using site-level cross-validation. This process involved training an AKI prediction model on one site (the source site) and testing it on the remaining sites (target sites), thereby allowing us to assess the transportability and adaptability of the models across health systems. Each site’s model was also evaluated on its own test set. This approach resulted in 81 internal/external evaluation combinations. After model training, a separate Shapley Additive Explanations (SHAP) library[20] was used to compute SHAP values for estimating the importance of predictors. SHAP values, expressed in log-odds, represent the individual contribution of each predictor to the prediction.

### 2.4 Univariate Meta-regression

To examine biomarker-specific dose–response relationships of common predictors on AKI risk across sites, univariate meta-analysis for the SHAP value of each predictor was performed with each site data treated as a separate study. We identified 30 predictors that most frequently appeared among the top 30 predictors across all site-level models for conducting univariate meta-regression. Univariate analyses involving continuous SHAP values were performed using a generalized additive model (GAM)[21]. Analogous to traditional logistic regression where log-odds are fitted linearly, we applied three fittings to the SHAP values, indicated as *φ*. The fitting formula takes the following form:

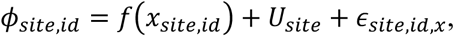

Here, *U_site_* represents the random effect terms arising from the underlying differences in patient distribution. *x* represents a predictor and *f*(*x*) represents the transformation of the predictor using linear, quadratic, and spline basis functions. *ε_site_*_,*id*,*x*_ represents the idiosyncratic error term.

We computed the coefficient of determination (*r*^2^) to assess the proportion of variance in SHAP values explained by the model. A higher *r*^2^ indicates that the predictor exhibits stable and robust predictive influence across multiple sites, thereby supporting its generalizability.

### 2.5 Bivariate Meta-regression

To evaluate how the joint effect of two predictors influences the risk of AKI, we conducted a bivariate meta-analysis. This approach enables the identification of predictor interactions that may improve predictive performance and uncover synergistic relationships between predictors overlooked by univariate analyses. By uncovering these interactions, we gain deeper insights into the pathophysiology of AKI and support more personalized approaches to patient management. The formula for this analysis is therefore modified as follows:

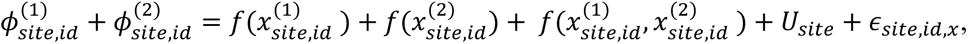

for continuous/continuous predictors, where we focus only on using cubic spline functions. For discrete predictors, the formula becomes:

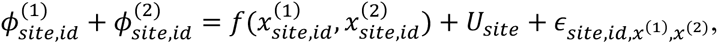

where *x*^(2)^ is a binary variable representing the contribution by *x*^(1)^ in the absence or presence of *x*^(2)^ separately. *φ*^(1)^ and *φ*^(2)^ represent the SHAP values of predictor 1 and 2.

We selected the top 15 most generalizable predictors based on *r*^2^ from the univariate meta-regressions. For each primary predictor, all possible predictor pairs were evaluated, and the improvement in *r*^2^ was calculated relative to the univariate models of the individual predictors.

## 3. Results

### 3.1 Performance of Site-Level Prediction Models Across Sites

Using patient-level data from nine sites within the GPC and PaTH networks (**Table 1**), we trained and cross-validated a CatBoost model at the site level. **Figure 1** illustrates the area under the receiver operating characteristic curve (AUROC) across 81 internal/external evaluation combinations, reflecting the model’s performance across sites. When evaluated on source site testing data (i.e., training and testing data originating from the same institution; as shown in the diagonal cells of the figure), the model’s AUROC ranged from 0.79±0.01 to 0.88±0.01. When evaluated on target sites, the AUROC ranged from 0.67±0.01 to 0.82±0.003. Model transportability, defined as the average AUROC of a model evaluated on target sites divided by the AUROC of the model evaluated on source site testing data, ranged from 0.76±0.14 to 0.94±0.13.

**Figure 1:**
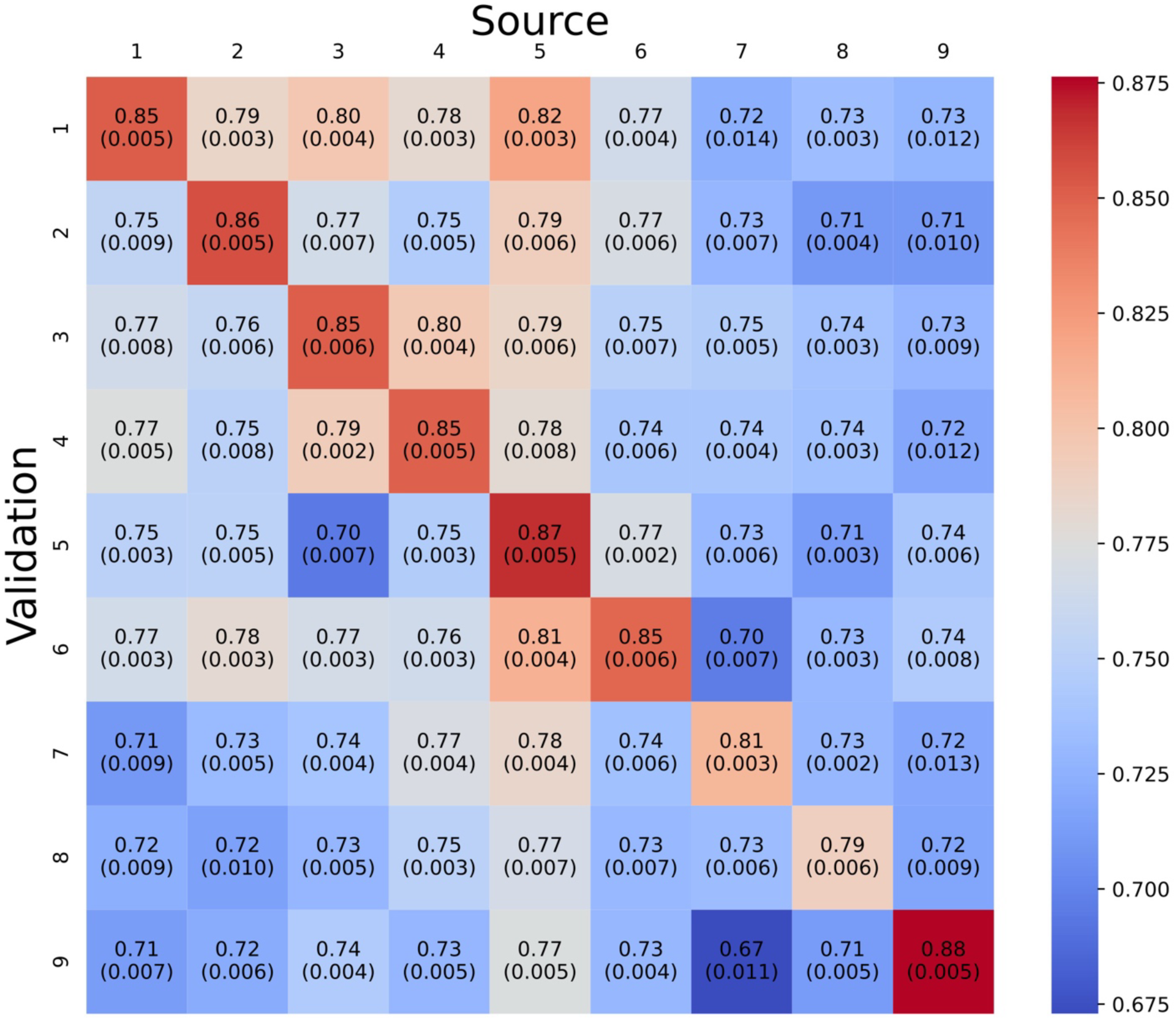
AUROC disparities across sites. This figure illustrates the average AUROC (standard deviation) for each site model applied to its own (source) as well as test data from other sites (target), represented as shaded areas. Each axis represents the data site. For instance, the leftmost column represents the model that was trained on site 1 training data (“Source”) and evaluated on test datasets (“Validation”) from site 1 and the other sites (2 to 9).

**Table 1.** Demographic characteristics at different health systems.

|  | 1 | 2 | 3 | 4 | 5 | 6 | 7 | 8 | 9 |
| --- | --- | --- | --- | --- | --- | --- | --- | --- | --- |
| **Sample number** |  |  |  |  |  |  |  |  |  |
| Encounters | 108,477 | 211,142 | 124,093 | 133,779 | 75,350 | 101,456 | 384,414 | 104,402 | 111,403 |
| Unique Patients | 57,246 | 106,479 | 69,399 | 82,256 | 47,799 | 59,553 | 223,949 | 69,064 | 69,752 |
| **AKI (encounter-level)** |  |  |  |  |  |  |  |  |  |
| Non-AKI | 92,583<br>(85.3%) | 182,642<br>(86.5%) | 107,585<br>(86.7%) | 115,904<br>(86.6%) | 64,575<br>(85.7%) | 87,438<br>(86.2%) | 331,690<br>(86.3%) | 87,495<br>(83.8%) | 100,473<br>(90.2%) |
| Any AKI | 15,894<br>(14.7%) | 28,500<br>(13.5%) | 16,508<br>(13.3%) | 17,875<br>(13.4%) | 10,775<br>(14.3%) | 14,018<br>(13.8%) | 52,724<br>(13.7%) | 16,907<br>(16.2%) | 10,930<br>(9.8%) |
| **Age (encounter-level)** |  |  |  |  |  |  |  |  |  |
| 18-25 | 4,308<br>(4.0%) | 8,990<br>(4.3%) | 4,826<br>(3.9%) | 5,638<br>(4.2%) | 4,004<br>(5.3%) | 4,150<br>(4.1%) | 9,423<br>(2.5%) | 7,359<br>(7.0%) | 5,864<br>(5.3%) |
| 26-35 | 8,601<br>(7.9%) | 17,092<br>(8.1%) | 9,749<br>(7.9%) | 10,311<br>(7.7%) | 6,641<br>(8.8%) | 8,485<br>(8.4%) | 19,679<br>(5.1%) | 13,275<br>(12.7%) | 11,711<br>(10.5%) |
| 36-45 | 10,497<br>(9.7%) | 20,457<br>(9.7%) | 10,601<br>(8.5%) | 13,131<br>(9.8%) | 7,896<br>(10.5%) | 9,929<br>(9.8%) | 24,441<br>(6.4%) | 14,854<br>(14.2%) | 12,952<br>(11.6%) |
| 46-55 | 15,410<br>(14.2%) | 32,942<br>(15.6%) | 18,511<br>(14.9%) | 22,846<br>(17.1%) | 12,360<br>(16.4%) | 15,187<br>(15.0%) | 41,413<br>(10.8%) | 22,083<br>(21.2%) | 16,738<br>(15.0%) |
| 56-65 | 23,512<br>(21.7%) | 49,655<br>(23.5%) | 28,207<br>(22.7%) | 31,969<br>(23.9%) | 16,468<br>(21.9%) | 22,495<br>(22.2%) | 75,550<br>(19.7%) | 24,853<br>(23.8%) | 24,358<br>(21.9%) |
| ≥ 66 | 46,149<br>(42.5%) | 82,006<br>(38.8%) | 52,199<br>(42.1%) | 49,884<br>(37.3%) | 27,981<br>(37.1%) | 41,210<br>(40.6%) | 213,908<br>(55.6%) | 21,978<br>(21.1%) | 39,780<br>(35.7%) |
| **Sex** |  |  |  |  |  |  |  |  |  |
| Male | 52,581<br>(48.5%) | 107,861<br>(51.1%) | 62,756<br>(50.6%) | 70,080<br>(52.4%) | 37,428<br>(49.7%) | 52,069<br>(51.3%) | 183,737<br>(47.8%) | 56,614<br>(54.2%) | 59,115<br>(53.1%) |
| Female | 55,895<br>(51.5%) | 103,279<br>(48.9%) | 61,337<br>(49.4%) | 63,698<br>(47.6%) | 37,922<br>(50.3%) | 49,385<br>(48.7%) | 200,677<br>(52.2%) | 47,724<br>(45.7%) | 52,287<br>(46.9%) |
| **Hispanic** |  |  |  |  |  |  |  |  |  |
| Yes | 12,661<br>(11.7%) | 11,701<br>(5.5%) | 3,931<br>(3.2%) | 3,334<br>(2.5%) | 1,234<br>(1.6%) | 5,193<br>(5.1%) | 1,880<br>(0.5%) | 65,621<br>(62.9%) | 9,044<br>(8.1%) |
| No | 90,592<br>(83.5%) | 197,965<br>(93.8%) | 119,815<br>(96.6%) | 124,916<br>(93.4%) | 71,316<br>(94.6%) | 95,947<br>(94.6%) | 353,666<br>(92.0%) | 38,051<br>(36.4%) | 98,461<br>(88.4%) |
| Unknown | 5,224<br>(4.8%) | 1,476<br>(0.7%) | 271<br>(0.2%) | 4,479<br>(3.3%) | 425<br>(0.6%) | 240<br>(0.2%) | 20,111<br>(5.2%) | 634<br>(0.6%) | 3,016<br>(2.7%) |
| **Race** |  |  |  |  |  |  |  |  |  |
| Asian | 2,275<br>(2.1%) | 1,964<br>(0.9%) | 1,370<br>(1.1%) | 1,198<br>(0.9%) | 340<br>(0.5%) | 928<br>(0.9%) | 2,339<br>(0.6%) | 1,321<br>(1.3%) | 1,588<br>(1.4%) |
| Black | 21,829<br>(20.1%) | 33,198<br>(15.7%) | 27,312<br>(22.0%) | 6,498<br>(4.9%) | 6,531<br>(8.7%) | 10,506<br>(10.4%) | 41,644<br>(10.8%) | 6,980<br>(6.7%) | 1,945<br>(1.7%) |
| Native American | 254<br>(0.2%) | 875<br>(0.4%) | 416<br>(0.3%) | 436<br>(0.3%) | 162<br>(0.2%) | 1,052<br>(1.0%) | 596<br>(0.2%) | 110<br>(0.1%) | 2,254<br>(2.0%) |
| White | 70,007<br>(64.5%) | 157,588<br>(74.6%) | 90,567<br>(73.0%) | 120,565<br>(90.1%) | 66,504<br>(88.3%) | 82,622<br>(81.4%) | 328,765<br>(85.5%) | 93,097<br>(89.2%) | 94,050<br>(84.4%) |
| Not specified | 5,284<br>(4.9%) | 16,320<br>(7.7%) | 2,434<br>(2.0%) | 123<br>(0.1%) | 883<br>(1.2%) | 4,301<br>(4.2%) | 186<br>(0.0%) | 106<br>(0.1%) | 9,994<br>(9.0%) |

Variation in model transportability across sites motivated us to investigate the underlying AKI risk predictors. As shown in **Figure 2**, their relative importance rankings highlight both shared and site-specific patterns. Several predictors, such as anion gaps 3 and 4, exhibited high importance at only a few sites, whereas others—including SCr, leukocytes, glucose, chloride, and age—consistently ranked among the most influential predictors across all models.

**Figure 2:**
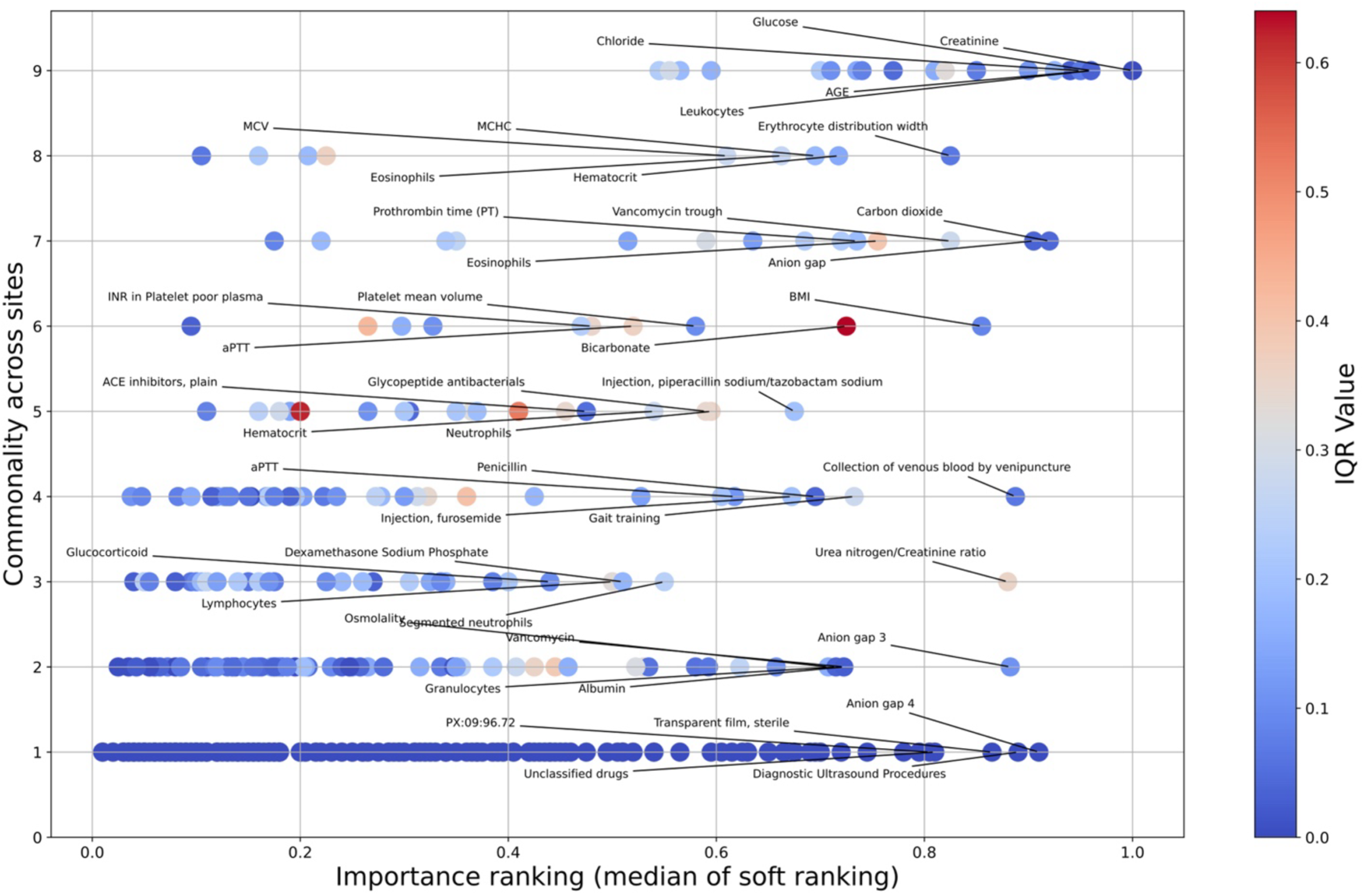
Predictor importance disparities across sites. This diagram illustrates the variability in predictor importance across site-level models. Each point represents a predictor that ranks within the top 100 in at least one of the nine site-level models. The y-axis indicates the frequency of sites recognizing the predictor as their top 100, termed as “site commonality.” The x-axis denotes the median importance ranking of the variables, referred to as “soft ranking”, where a value closer to 1 signifies higher importance. Additionally, the variation in ranking across sites, indicated by the interquartile range (IQR), is represented by the color intensity of each dot, with a higher IQR signifying greater discrepancies among sites.

### 3.2 Impact of Common Predictors on AKI Risk

SHAP values were calculated for each model based on its original training site and across other sites, as previously illustrated in **Figure 1**. The 8 best-fitting features, as determined by *r*^2^, are shown in **Figure 3**, while additional features are presented in **Figures S2**–**S3**. Detailed fitting results are in **Tables S3**–**S5**.

**Figure 3:**
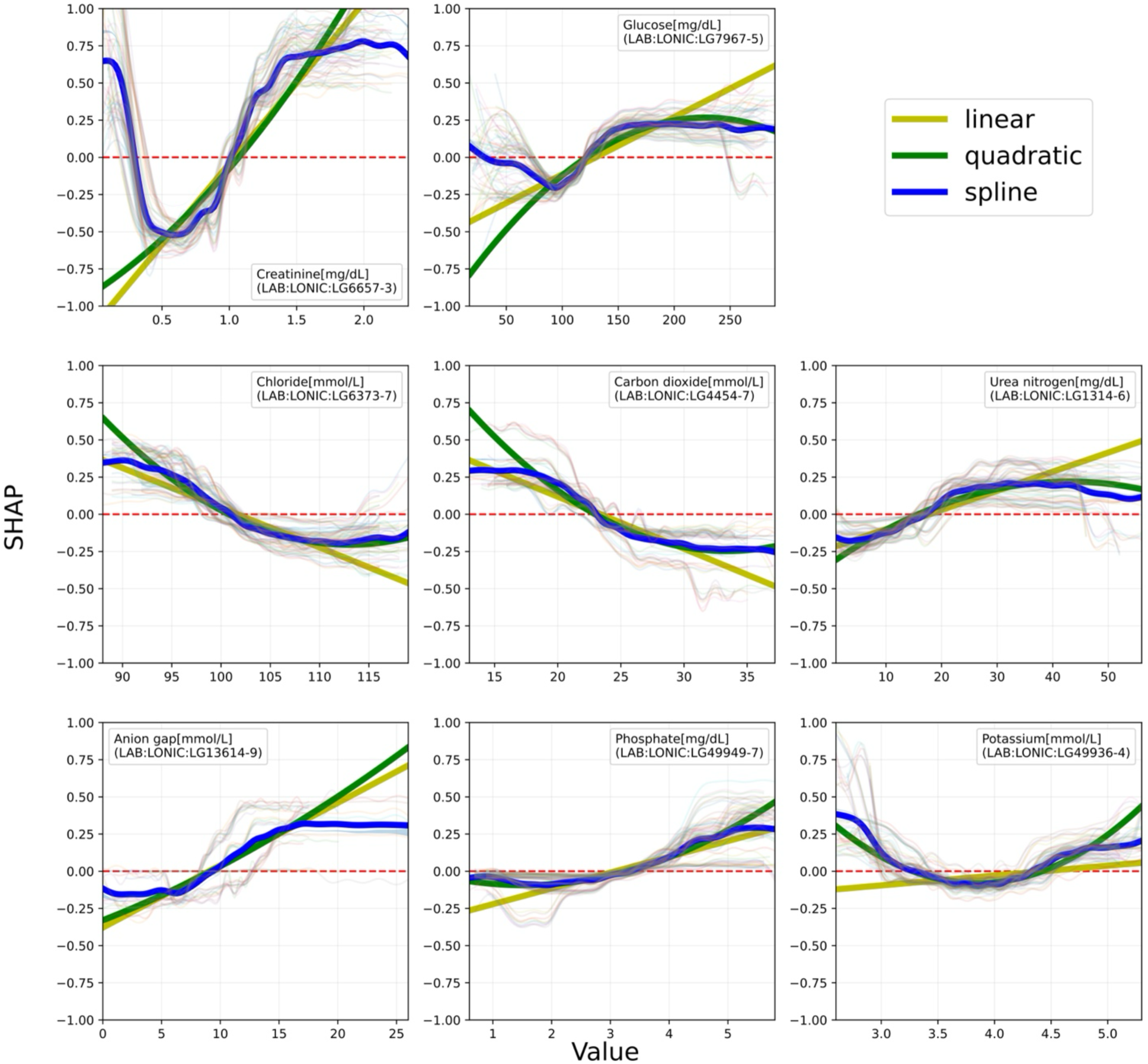
Meta-regression results for top continuous predictors. The x-axis represents the values for each predictor, spanning the range between the 5th and 95th percentiles of all values in the dataset. The y-axis represents SHAP values for that predictor. The light, background lines correspond to all 81 individual source/validation pairs. The bold lines (olive, green or blue) represent the meta-regression curves, fitted by three different methods. All variable values are collected as the most recent value 24-hr prior to the anchored event (onset for AKI patients; last SCr measurement day for non-AKI patients).

As shown in **Figure 3**, some predictors were adequately represented by linear or sigmoid-like functions, such as phosphate. However, potassium exhibited a quadratic relationship, indicating that both low and high values are associated with an increased risk of AKI. This relationship translates to a risk increase of approximately exp(0.3) to exp(0.38), corresponding to a 1.35- to 1.46-fold difference between the minimum and maximum risk. Similar patterns were observed for calcium and sodium (**Figures S2** and **S3**). Additional causal analysis results regarding the ranges of these ionic compounds are presented in **Supplementary Methods S1** and **S2**, **Figures S4**–**S6**, and **Table S6**. SCr showed anomalously higher AKI risk at very low values, deviating from linear or quadratic patterns and best captured using spline modeling. A sharp escalation in AKI risk was observed for glucose levels between 100 and 140 mg/dL, slightly above the normal fasting glucose threshold. This corresponded to an approximate exp(0.38) ∼ 1.46-fold increase in risk. In contrast, AKI risk remained near baseline levels around 70 mg/dL, within the hypoglycemic region. Both chloride and anion gap were associated with increased AKI risk within their reference ranges. Chloride levels between 96 and 100 mEq/L, within the reference range, were associated with an increased predicted risk of AKI, corresponding to an odds ratio (OR) of 1.28 (log-odds increase ∼0.25). Below 96 mEq/L, in the hypochloremia range, chloride levels were associated with a greater risk of AKI. Anion gap showed a linear increase around the reference range and flattened at both ends. Within the range of 6–16 mmol/L, an approximately exp(0.43) ∼ 1.54-fold increase in AKI risk was observed, while within the reference range of 4–12 mmol/L, the increase was approximately 1.14-fold. The most important continuous predictor in the meta-regression is SCr. At the highest values, it correlates with approximately exp(1.25) ∼ 3.5-fold difference, while at the lowest values, it correlates with exp(1.2) ∼ 3.3-fold difference, between minimum and maximum risk.

### 3.4 Predictor Generalizability Across Sites

To evaluate the consistency of predictor effects across distinct health systems, we derived the *r*^2^ value of the meta-analysis fit, as shown in **Figure S7**. Various predictors exhibited differing marginal effects across the target and source models, and the degree of agreement between these models influenced the resulting meta-regression *r*^2^. For instance, SCr, glucose, chloride, carbon dioxide, and BUN demonstrated a high degree of functional similarity across sites, indicating a consistent functional relationship between the values of these variables and the predicted risk of AKI. This consistency also suggests superior and more uniform data quality for these predictors across distinct health systems. Additionally, SCr, glucose, chloride, and carbon dioxide are also highly important predictors across sites, as indicated by their commonality across site-level models (**Figure 2** and **Figure S8**). Conversely, leukocyte was important in most models, but exhibited high functional heterogeneity across sites, leading to poor curve fitting in meta-regression.

### 3.5 Interactions Among Top Predictors

To evaluate how joint effects of two predictors influence AKI risk, we conducted a bivariate meta-analysis. This approach enables the identification of predictor interactions that improve AKI predictive performance and uncover synergistic relationships between predictors overlooked by univariate analyses. Detailed fitting results are presented in **Tables S7** and **S8**.

For continuous-to-continuous bivariate meta-regression, the top 10 predictor pairs that show the greatest improvement in *r*^2^ compared to the univariate curves of both predictors are presented in **Figure 4**. The interaction terms and contour plots of the interactions are presented in **Figures S9** and **S10**. Systolic blood pressure demonstrated a significant interaction with erythrocyte distribution width (EDW) when ≥140 mmHg. Age was also found to significantly interact with EDW. SCr was found to significantly interact with BUN when SCr ≥1.4 mg/dL.

**Figure 4:**
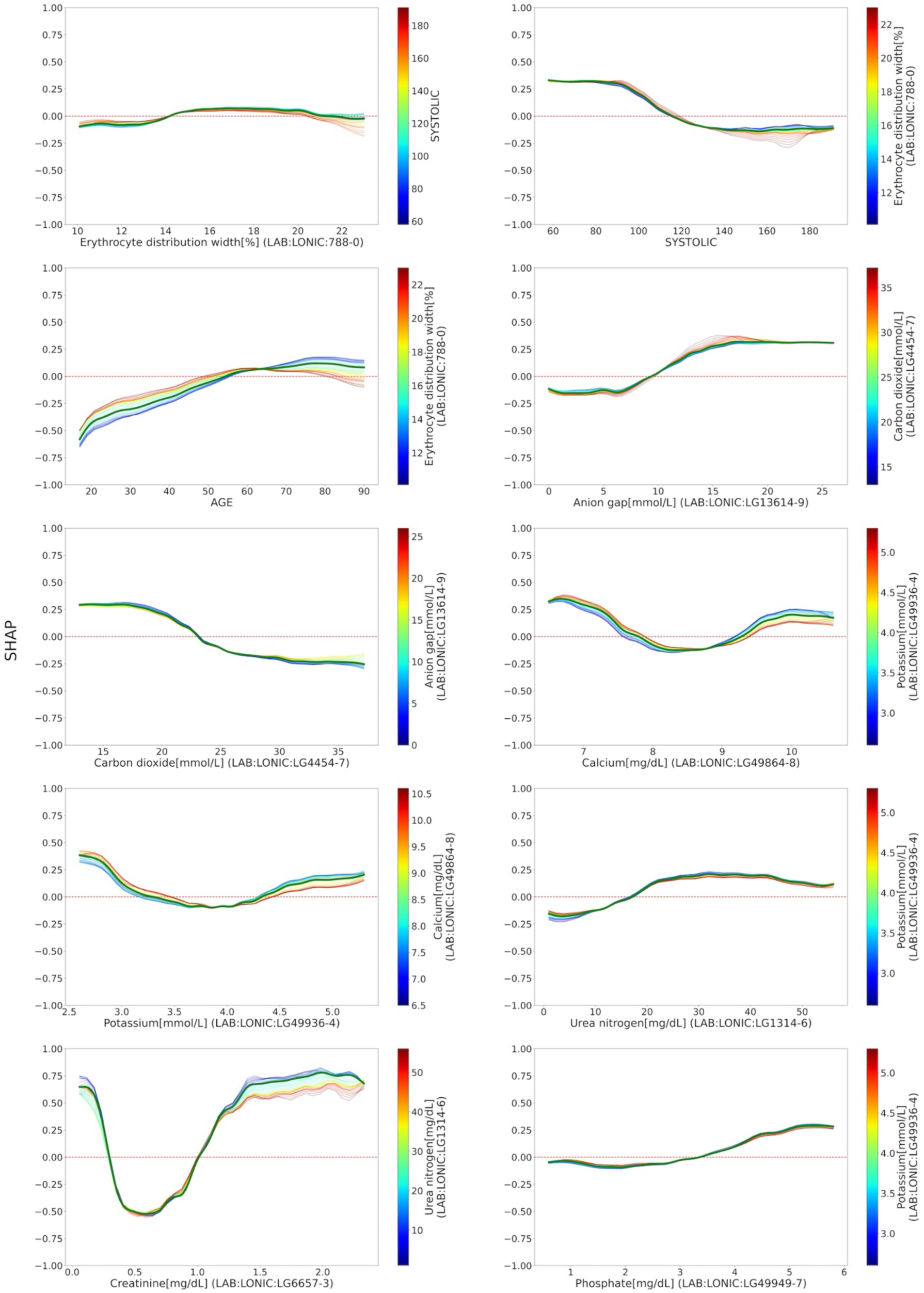
Continuous-continuous bivariate analysis. This figure illustrates the interaction between two continuous predictors. It highlights pairs with the greatest improvement in *r*^2^ compared to the maximum of univariate *r*^2^ of the two predictors. The green curve shows the univariate SHAP value for the predictor on the x-axis, while the profile lines represent the SHAP values from the bivariate interaction at constant value of the second predictor, as indicated by the color bar.

Additionally, the top 15 predictor pairs where the secondary predictor showed the greatest improvement in *r*^2^ compared to univariate regression of the primary predictor are shown in **Figures S11–13**. **Figure S11** highlights that SCr, the most important primary predictor, also emerged as the most significant secondary predictor in improving *r*^2^. In addition to the interactions that yielded the greatest improvements in *r*² for both predictors, SCr also significantly interacted with eosinophils, EDW, vancomycin trough levels, age, and chloride, each contributing to improvements in the *r*² of the primary predictor, although the patterns of interaction differed across variables.

For the binary-to-continuous bivariate meta-regression, the analysis focused on the only binary predictor among the top 15 best predictors—collection of venous blood by venipuncture (PX:CH:36415). The results are presented in **Figures S14** and **S15**. Overall, all observed interactions exhibited approximately a 10% difference in AKI risk.

## Discussion

In this study, we evaluated machine learning-based AKI prediction models across nine academic medical centers, conducting a total of 81 performance evaluations. Our findings revealed significant variability in predictor importance and marginal effects across sites, which contributed to performance declines in external validation. To identify robust and generalizable risk predictors, we implemented meta-regressions to systematically quantify the marginal effects of individual predictors across site-specific models.

Even among highly predictive AKI predictors, some exhibited greater consistency across sites, while others showed notable variability. This variability can be attributed to several factors. Firstly, differences in the frequency of measurements taken at different sites can affect how current the recorded values are for the prediction. Secondly, the predictive performance of certain predictors can vary across sites due to their sensitivity to population characteristics, such as demographics. Some clinical lab tests are known to exhibit racial or ethnic-specific differences, which can influence their values and interpretation across different patient populations. Finally, discrepancies can also arise from variations in measurement methods, including differences in lab equipment and techniques used across sites. This variation will be explored in more detail in the following paragraph. While such inconsistencies cannot be entirely resolved, our analysis identified several predictors that remain reliable across health systems, ensuring that AKI risk assessment can be applied broadly and accurately.

Some predictors reaffirmed established knowledge, such as SCr and BUN, which consistently emerged as the most important predictors of AKI.[17] Others are plausibly related to renal function such as serum electrolyte concentrations. One of our key findings was the identification of glucose as a significant and generalizable predictor. While previous studies reported a 1.48-fold increase in AKI risk per 100 mg/mL increase in glucose levels (log-odd = 0.39),[22] we observed a sharp escalation in AKI risk for glucose levels between 100-140 mg/mL, suggesting that even modest elevation of glucose levels can disproportionately increase AKI risk. This underscores the need for more frequent glucose monitoring, even at levels previously considered less concerning. **Figure S16** illustrates the relationship between glucose SHAP values and A1c levels, revealing smaller differences between diabetic and non-diabetic patients (log-odds = 0.20) excluding the prediabetic range. This suggests that short-term glucose variations may play a pivotal role in AKI risk.[23]

Chloride also emerged as an important and generalizable predictor of AKI. Our analysis indicated an increase in AKI risk within the reference range of chloride levels (between 96 and 100 mEq/L). In the hypochloremia region, the predicted risk of AKI was greater. This observation is consistent with a previous study, in which a multivariable logistic regression model identified a baseline serum chloride level of ≤94 mmol/L as independently associated with an increased risk of AKI.[24] Similarly, the anion gap demonstrated a linear increase in log-odds by approximately 0.43 within the range of 6–16 mmol/L (i.e., 1.54-fold increase in AKI risk), with risk saturation beyond this range. Even within the reference range of 4–12 mmol/L, there was a 1.14-fold increase in AKI risk. These findings indicate variations in predictive value of normal range in both chloride and anion gap, suggesting a potential need for vigilant monitoring in hospitalized patients.

Electrolytes such as calcium, potassium and sodium exhibited non-linear, quadratic relationships with AKI risk, with both low and high levels associated with increased risk. Interrupted time-series analyses identified specific medications as potential drivers of these non-monotonic relationships (**Figures S5** and **S6**). For example, bisphosphonates such as Pamidronate and Zoledronate were linked to a drop in calcium slope and an increase in SCr slope, potentially explaining the association between low calcium and AKI. Diarrhea medications like diphenoxylate and loperamide increased the slopes of both potassium and SCr, while Renin-angiotensin-aldosterone system blockers (e.g., eplerenone) elevated potassium levels, exacerbating AKI risk. Loop and thiazide diuretics reduced sodium levels and increased AKI risk, as evidenced by a negative change in sodium slope and a positive change in SCr. These findings align with previous studies, suggesting a potential causal role of medications in the electrolyte-AKI relationships.[25–28]

Leukocyte count showed significant site-level variability despite prior studies identifying leukocytosis as a marker for AKI.[25, 27, 29] This highlights the challenge of generalizing risk models across diverse clinical settings. A strong correlation between carbon dioxide and AKI was also observed, consistent with prior studies linking low bicarbonate levels to sepsis and shock, both of which can cause AKI.[18, 30] This suggests that low bicarbonate levels may act as a confounder in this patient group, warranting further investigation. Further analysis identified LDH, AST and uric acid – biomarkers of hemolysis and tumor lysis syndrome – as potential confounders for the association between electrolytes and AKI. However, these factors did not fully explain the non-linear correlations, necessitating further investigation into the underlying mechanisms.

Beyond single-variable effects, bivariate meta-regression revealed interactions between predictors that significantly altered AKI risk compared with univariate effects alone (**Figures 4** and **Figures S9**–**S15**). These interactions highlight pathways through which combinations of biomarkers jointly modulate AKI susceptibility.

A key limitation of our study is its reliance on observational associations rather than causal inference. Although interrupted time-series analysis and panel cointegration tests were used to explore causality in selected predictors, comprehensive causal inference remains an unmet need. Future research should prioritize causal approaches to refine predictive models and inform actionable interventions for AKI prevention.

In summary, our large, multi-center evaluation of ML-based AKI models identified robust and generalizable risk predictors across diverse healthcare systems. Meta-regression analyses provided clinical insights into value-specific risk drivers and nonlinear interactions, including glucose, calcium, potassium, and sodium. These findings enhance mechanistic understanding of AKI risk and provide a scalable framework for precision risk stratification and prevention in hospitalized patients.

## Data Availability

The data used in this study were de-identified EHRs from the Greater Plains Collaborative (GPC) and PaTH clinical research networks, that is not publicly available due to institutional and regulatory restrictions. Access to the data is governed by GPC and PaTH coordinating centers and requires institutional data use agreements and approval to protect patient privacy and compliance with ethical and legal considerations. Researchers interested in accessing similar data may contact GPC and PaTH front doors to inquire about potential data access.

## Data sharing statement

The data used in this study were de-identified EHRs from the Greater Plains Collaborative (GPC) and PaTH clinical research networks, that is not publicly available due to institutional and regulatory restrictions. Access to the data is governed by GPC and PaTH coordinating centers and requires institutional data use agreements and approval to protect patient privacy and compliance with ethical and legal considerations. Researchers interested in accessing similar data may contact GPC and PaTH front doors to inquire about potential data access. A GitHub repository containing the code used in the analysis is available at https://github.com/GatorAIM/AKI_MetaAnalysis.

## Generative AI Statement

During the preparation of this work, the authors used generative artificial intelligence tools solely for language editing and stylistic refinement. After using this tool/service, the authors reviewed and edited the content as needed and take full responsibility for the content of the publication.

## Authors’ contributions

ML initiated the project and the collaboration. ML, AY, and HYC designed the overall study. HYC extracted the study cohort and DL and QX contributed to data processing. HYC, DL, and QX performed experiments and data analysis. AY, JAK, and DYF contributed their clinical expertise in analyzing results. HYC, DL, and ML drafted and revised the manuscript with critical advice provided by AY, JAK, DYF, EAC, LGC, SC, AJA, JK, KMM, AM, BT, MS, LRW, and YH. DL, QX, and ML verified the underlying data reported in the manuscript.

## Appendix A. Supplementary data

**Supplementary Method S1**: Causal analysis of electrolytes.

**Supplementary Method S2**: Descriptions of interrupted time-series, cointegration, and confounding analyses.

**Figure S1**: Flowchart for estimating baseline SCr.

**Figure S2**: Meta-regression results for top features.

**Figure S3**: Meta-regression results for top features (cont.).

**Figure S4**: Lagged Pearson correlation analysis.

**Figure S5**: The change of slope interrupted time-series analysis of different medications.

**Figure S6**: The change of slope interrupted time-series analysis of different medication (cont.).

**Figure S7**: Functional similarity of predictors across sites.

**Figure S8**: Feature selection disparities across site.

**Figure S9**: Continuous-continuous bivariate analysis 1.

**Figure S10**: Continuous-continuous bivariate contour maps.

**Figure S11**: Continuous-continuous bivariate analysis 2.

**Figure S12**: Continuous-continuous bivariate analysis 3.

**Figure S13**: Continuous-continuous bivariate contour maps.

**Figure S14**: Discrete-continuous bivariate analysis.

**Figure S15**: Discrete-continuous bivariate analysis (cont.).

**Figure S16**: SHAP value of glucose vs corresponding A1c.

**Table S1**: Details of the medical codes in this study.

**Table S2**: Number of predictors used in final model per site.

**Table S3**: Linear fitting results.

**Table S4**: Quadratic fitting results.

**Table S5**: Spline fitting results.

**Table S6**: Results from Confounding Analysis.

**Table S7**: Fitting characteristics (Interaction).

**Table S8**: Fitting characteristics (Interaction) [Collection of venous blood by venipuncture].

