## Supplementary Materials for "Cross-System Meta-Analysis of Machine Learning Predictors Identifies Value-Specific Risk Drivers and Interactions Underlying Acute Kidney Injury"

### Contents of Supplementary Materials

**Supplementary Method S1**: Causal analysis of electrolytes.
**Supplementary Method S2**: Descriptions of interrupted time-series, cointegration, and confounding analyses.
**Figure S1**: Flowchart for estimating baseline SCr.
**Figure S2**: Meta-regression results for top features.
**Figure S3**: Meta-regression results for top features (cont.).
**Figure S4**: Lagged Pearson correlation analysis.
**Figure S5**: The change of slope interrupted time-series analysis of different medications.
**Figure S6**: The change of slope interrupted time-series analysis of different medication (cont.).
**Figure S7**: Functional similarity of predictors across sites.
**Figure S8**: Feature selection disparities across site.
**Figure S9**: Continuous-continuous bivariate analysis 1.
**Figure S10**: Continuous-continuous bivariate contour maps.
**Figure S11**: Continuous-continuous bivariate analysis 2.
**Figure S12**: Continuous-continuous bivariate analysis 3.
**Figure S13**: Continuous-continuous bivariate contour maps.
**Figure S14**: Discrete-continuous bivariate analysis.
**Figure S15**: Discrete-continuous bivariate analysis (cont.).
**Figure S16**: SHAP value of glucose vs corresponding A1c.
**Table S1**: Details of the medical codes in this study.
**Table S2**: Number of predictors used in final model per site.
**Table S3**: Linear fitting results.
**Table S4**: Quadratic fitting results.
**Table S5**: Spline fitting results.
**Table S6**: Results from Confounding Analysis.
**Table S7**: Fitting characteristics (Interaction).
**Table S8**: Fitting characteristics (Interaction) [Collection of venous blood by venipuncture].

### Supplementary Method S1: Causal analysis of electrolytes.

To explore the non-linear relationship between three key electrolytes (calcium, potassium, and sodium and the risk of AKI, we investigated specific medications and laboratory measurements as potential confounders of the association between the levels of these electrolytes and serum creatinine. For medications, interrupted time-series analysis (**Supplementary Method S2**) was utilized to assess the potential causal links between the administration of certain drugs and changes in electrolyte and serum creatinine levels. For laboratory measurements, panel co-integration tests and lagged Pearson correlation analyses were employed to identify causal roles for the potential confounding variables.

As an initial step, we used expert medical knowledge to hypothesize a set of potential confounding factors for each electrolyte, focusing on medications (like renin-angiotensin-aldosterone system, RAAS blockers, diuretics, and anti-diarrheal medications), and clinical/laboratory measurements indicative of conditions like hypotension or cell lysis syndromes. These medications and conditions are known to induce changes in electrolytes and could also potentially cause AKI. We also examined the impact of cell lysis syndromes (rhabdomyolysis, hemolysis, tumor lysis) as confounders by considering the effect of creatine kinase, AST, LDH, uric acid and potassium levels. Lagged Pearson correlation analysis indicates significant temporal correlations between AST, uric acid, potassium, and creatinine, suggesting potential causal relationships. In **Figure S4**, we show the lagged Pearson coefficients, where a significant maximal value in the positive lag means a potential causation of the cofounder to the ions and serum creatinine. Of all the combinations we tested, AST, LDH and uric acid show significant maximal correlation in the positive lagged regime, which are the confounders we consider in the subsequent analysis.

To examine the confounding effect of medications, we examined a list of eight medications with different potential confounding mechanisms. For instance, bisphosphonates are often used to treat hypercalcemia and so are associated with high calcium levels, but AKI is a potential adverse reaction. Loop and thiazide diuretics cause hyponatremia and hypokalemia. They are given frequently to patients with congestive heart failure, who are predisposed to AKI, and can also directly cause AKI. Conversely, the aquaretic class of diuretics is indicated for the treatment of hyponatremia but can cause AKI.  Anti-diarrheal medications are often prescribed to patients with diarrhea, which can be a cause of low potassium level and AKI. Inhibitors of the renin-angiotensin-aldosterone system, including angiotensin II receptor blocker, renin-inhibitors and Raldosterone antagonists can cause high potassium level and also cause AKI.

To test these potential causal mechanisms, we applied interrupted time-series tests. **Figures S5** and **S6** show the forest plot of the difference between post-intervention slope and pre-intervention slope, indicating the change of trend of the time series before and after initiating treatment with the medication. Bisphosphonates, antidiarrheals, aldosterone antagonists, loop diuretics and thiazides significantly altered the trend of the time series for both serum creatinine and the corresponding electrolyte concentrations. All five medication classes increased the trend of creatinine, thus increasing risk of or causing AKI. Medication associated due to treatment of the corresponding ion imbalance (Bisphosphonates, Antipropulsives, and Aquaretic) could reduce or reverse the trend of the high ionic imbalance, but could cause low imbalance, which both are associated with high risk of AKI, while Medication associated with the cause (Aldosterone antagonists and Diuretics) enhances the imbalance.

Following the identification of these confounders, a multivariate linear regression analysis was conducted to assess their overall impact. AST, LDH, Uric acid and the five medications identified as having potential causal relationships with both serum creatinine and one of the electrolyte levels were included as covariates in the adjusted model. The result is shown in **Table S6**, where $\beta$ is the unadjusted slope and $\beta'$ is the adjusted slope. $\beta'$ represents the change in the outcome variable for a one-unit change in the predictor variable. In an unadjusted model, $\beta$ reflects the direct relationship between the predictor and outcome, without accounting for other variables. In an adjusted model, $\beta'$ reflects the relationship between the predictor and outcome after controlling for other covariates previously identified, isolating the effect of the predictor on the outcome. Although the presence of confounding effects was noted as the change in slope is more than 10%, they generally do not completely account for the causal relation. This result suggests a direct causal link between serum creatinine levels and the electrolytes or the presence of unknown confounders, in the high potassium low and high calcium region where the adjusted slope is significantly different from zero and follow the U-shape trend in their respective region.

### Supplementary Method S2: Descriptions of interrupted time-series, cointegration, and confounding analyses.

**Interrupted time-series analysis.** The general formula for the regression is

$$y_{t,id} = \beta_0 + \beta_1 t + (\Delta\beta_0 + \Delta\beta_1 t)D_{id} + \epsilon_{id},$$

where $y$ is the value for SCr/potassium/calcium/sodium, $t$ is the time component, $T$ is the dummy variable, with $D = 0$ for pre-treatment data and $D = 1$ for post-treatment data, $\beta_0$ and $\Delta\beta_0$ indicates the intercept and its variation with $\beta_1$ and $\Delta\beta_1$ representing the slope and its variation. Individual patients were fitted, and the value of $\Delta\beta_0$ and $\Delta\beta_1$ was evaluated by *t*-test.

**Panel cointegration test**. We performed an *Engle-Granger two-step cointegration test*[1] on panel data with random effects at the patient level. First, a panel unit roots test was performed to show the existence of unit root, then a one-way random effect model was fitted between the targeted lab measurement and calcium, potassium, sodium or SCr, with the functional form:

$$y_{t,id} = \beta_0 + \beta_1 x_{t,id} + U_{id} + \epsilon_{id,x},$$

where $y$ is the value for SCr/potassium/calcium/sodium, $x$ represents the target lab measurement and $U_{id}$ corresponds to nested random effect on site level and encounter level. The residual was tested for a unit root using *Augmented Dickey–Fuller test*[2].

**Confounding variable analysis** We performed a multivariate linear regression[2] on calcium, potassium, and sodium against the  value. For continuous confounders, we compared the slope between the regression with and without the cofounding variable using the following formula:

$$y = \alpha + \beta x + \epsilon_x, \quad (1)$$

$$y = \alpha' + \beta' x + \beta_z z + \epsilon_{x,z}, \quad (2)$$

where $y$ is the SCr value, $x$ is the calcium, potassium or sodium value and $z$ is the corresponding confounder. All values are standardized. We considered the confounding effect significant if change in $\beta$ was more than 10% of $\beta'$. In this analysis, we restrict the sample to be within the respective range of patients as identified in **Figure 3**.

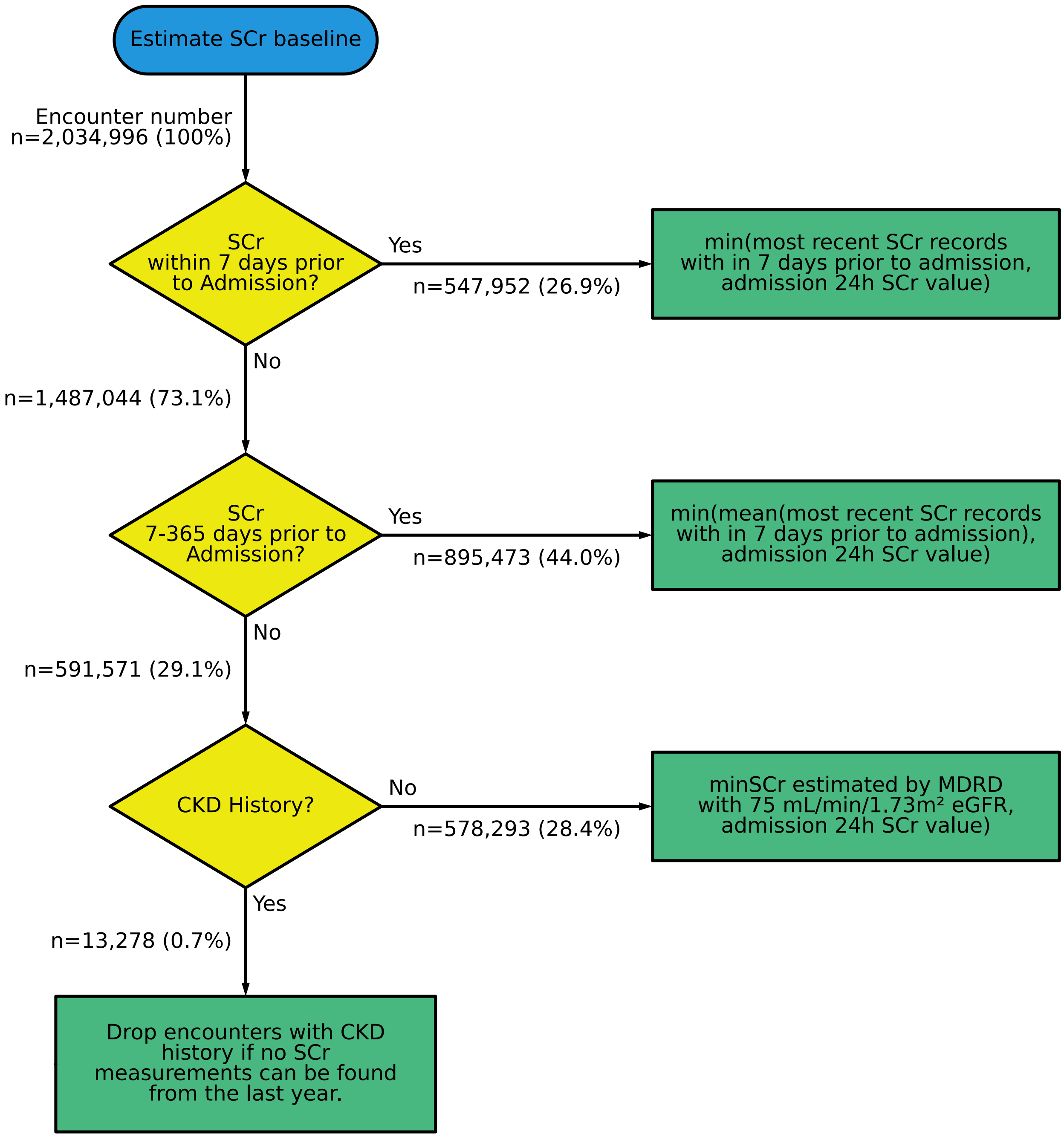


**Figure S1: Flowchart for estimating baseline SCr**.

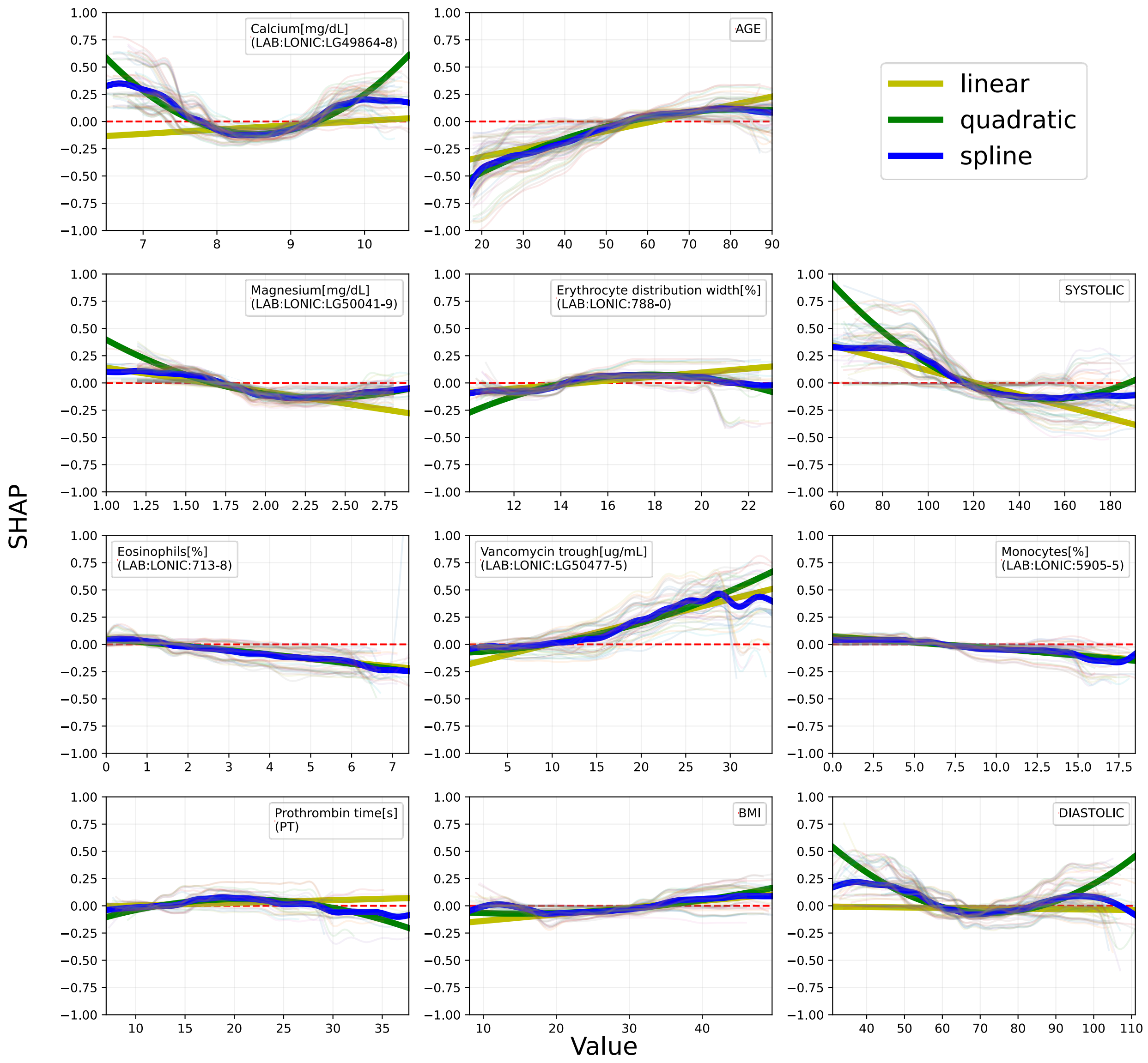


**Figure S2: Meta-regression results for top features**. The bold lines (olive, green or blue) represent the meta-regression curves, while the light, background lines correspond to individual source/validation pairs.

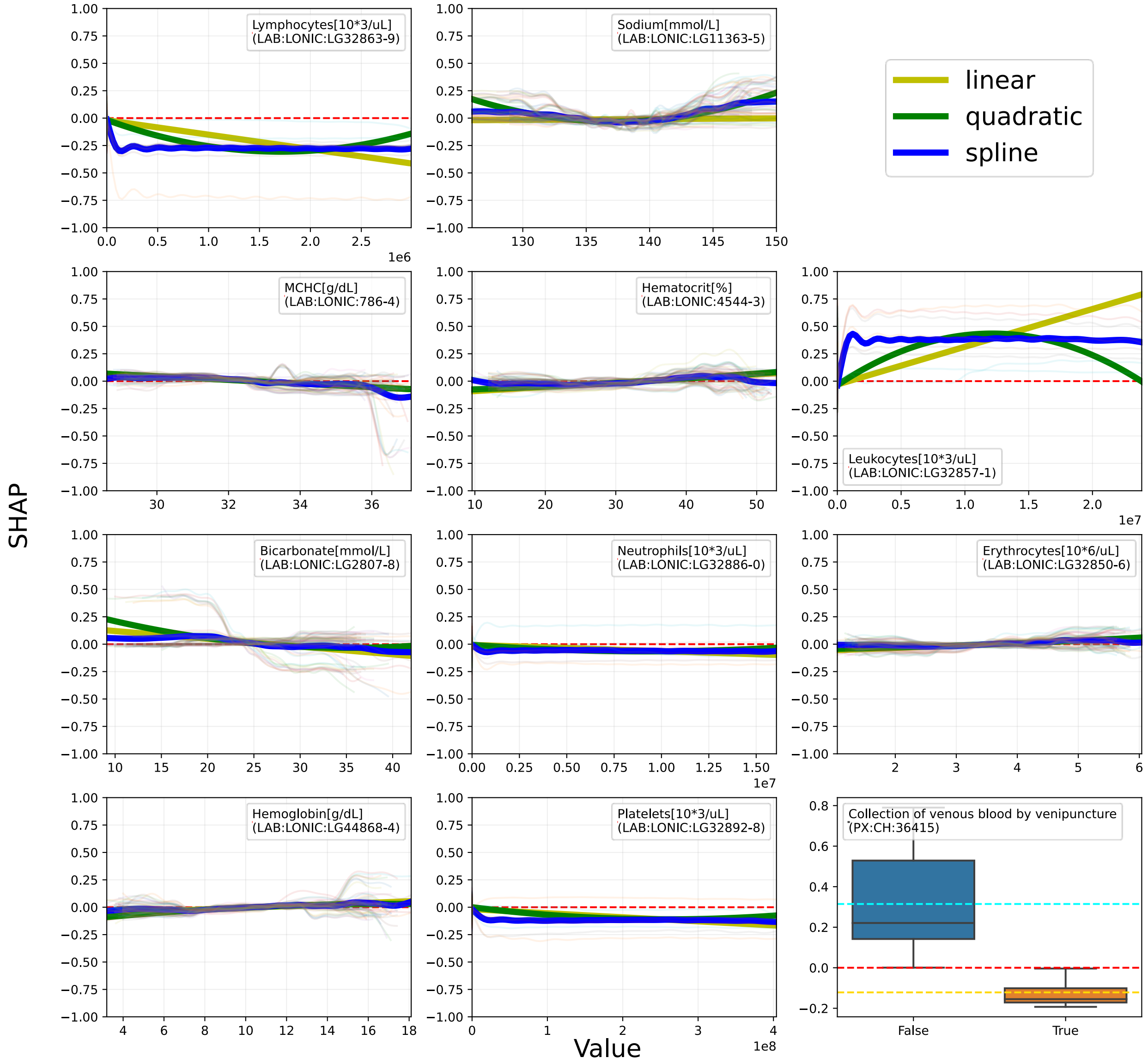


**Figure S3: Meta-regression results for top features**. (cont.) The bold lines (olive, green or blue) represent the meta-regression curves, while the light, background lines correspond to individual source/validation pairs.

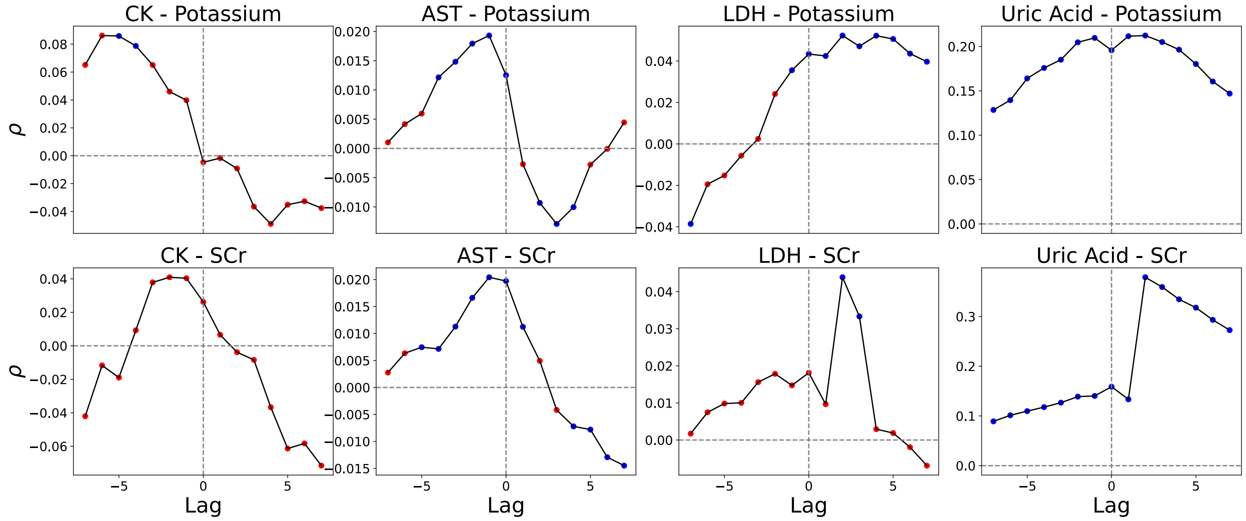


**Figure S4: Lagged Pearson correlation analysis**. The time series for each potential confounder is compared to the future/past time series of potassium and serum creatinine (SCr) shifted by “Lag” number of days. ρ is the Pearson correlation coefficient. A positive lag means the confounder time series is being compared to the future value of potassium or SCr. Significant values are denoted by blue dots, while non-significant values are denoted by red dots. A significant value in the positive lag region indicates a potential correlation between the past value of the confounder and the future value of potassium/SCr, suggesting a potential causal relationship.

| label | HR (95% CI) | p-val |
|---|---|---|
| Aldosterone antagonists(HK):C03DA | | |
| potassium | 0.05 (0.05–0.05) | 0.00 |
| sCr | 0.02 (0.02–0.02) | 0.00 |
| Angiotensin II receptor blockers (ARBs)(LNa):C09DX | | |
| sodium | -0.03 (-0.12–0.06) | 0.53 |
| sCr | 0.02 (0.02–0.03) | 0.00 |
| Antipropulsives(LK):A07DA | | |
| potassium | 0.02 (0.02–0.02) | 0.00 |
| sCr | 0.01 (0.01–0.01) | 0.00 |
| Bisphosphonates(HCa):M05BA | | |
| calcium | -0.05 (-0.07–-0.04) | 0.00 |
| sCr | 0.01 (0.01–0.01) | 0.00 |
| Sulfonamides, plain(LKLNa):C03CA | | |
| sodium | -0.29 (-0.30–-0.29) | 0.00 |
| sCr | 0.01 (0.01–0.01) | 0.00 |
| Sulfonamides, plain(LNa):C03BA | | |
| sodium | -0.16 (-0.19–-0.13) | 0.00 |
| sCr | 0.02 (0.01–0.02) | 0.00 |
| Thiazides, plain(LNa):C03AA | | |
| sodium | -0.22 (-0.24–-0.20) | 0.00 |
| sCr | 0.01 (0.00–0.01) | 0.00 |

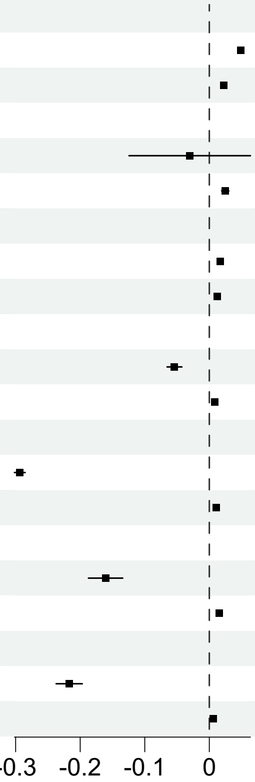


**Figure S5: The change of slope interrupted time-series analysis of different medications.** The x-axis represents the difference in slope of the laboratory measurement per unit time in days between the time period post-intervention with the indicated medication classes and pre-intervention.

| label | HR (95% CI) | p-val |
|---|---|---|
| Renin-inhibitors(LNa):C09XA | | |
| sodium | -0.11 (-0.57–0.36) | 0.66 |
| sCr | -0.02 (-0.04–0.01) | 0.20 |
| Vasopressin antagonists(LNa):C03XA | | |
| sodium | 1.80 (1.66–1.93) | 0.00 |
| sCr | 0.01 (0.00–0.01) | 0.01 |

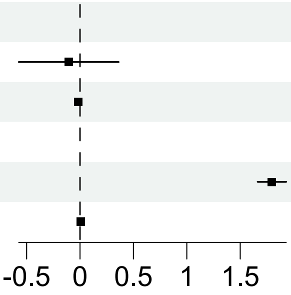


**Figure S6: The change of slope interrupted time-series analysis of different medication (cont.).** Figures were split due to differences in the x-axis scaling.

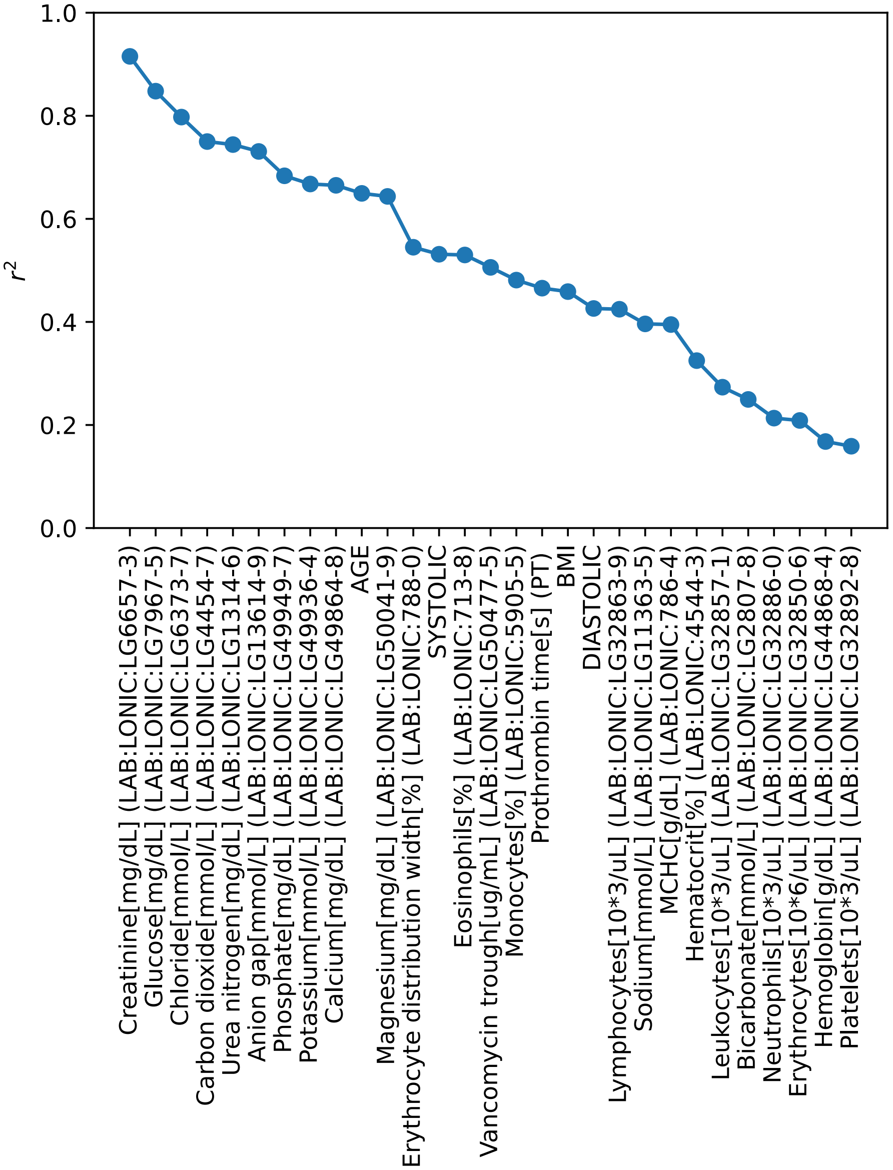


**Figure S7: Functional similarity of predictors across sites.** This figure illustrates the $r^2$ value of the spline fit for top continuous predictors.

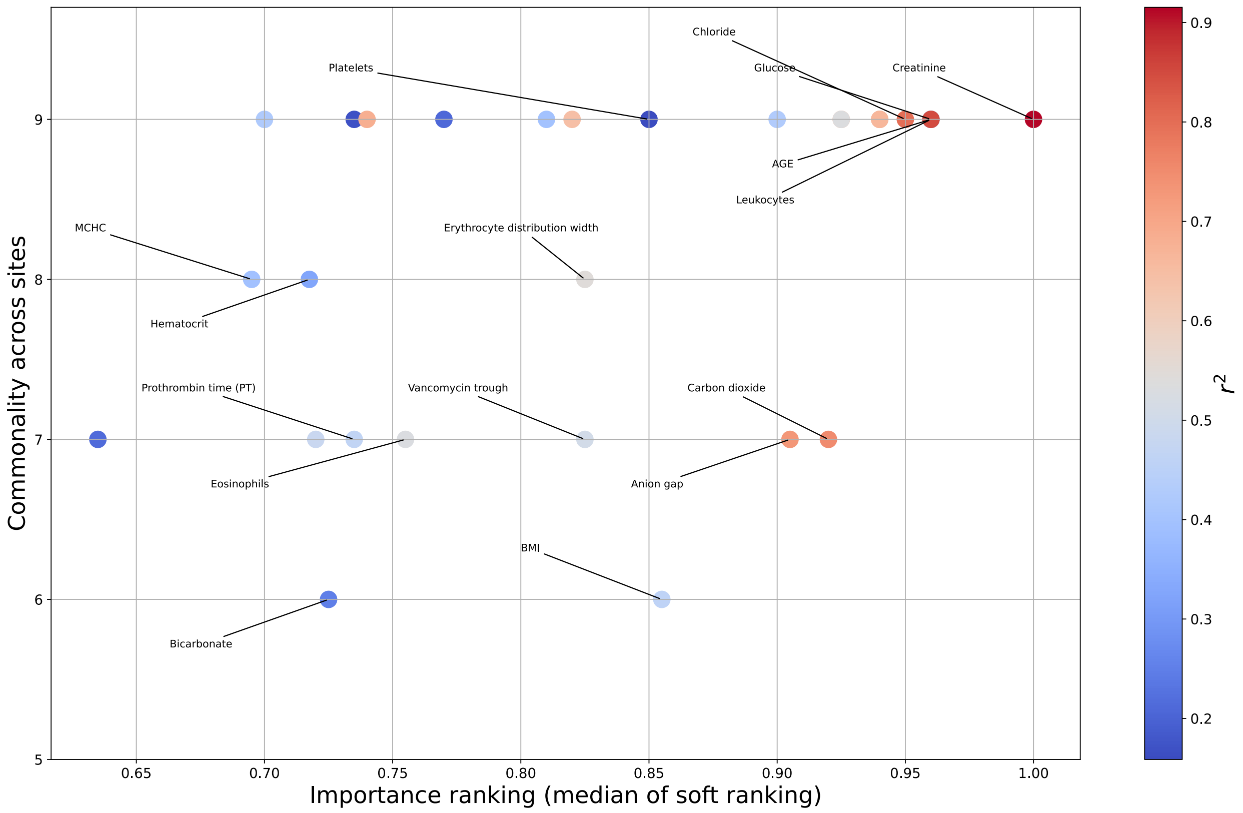


**Figure S8: Feature selection disparities across sites.** Color represents the $r^2$ value from **Figure S7**.

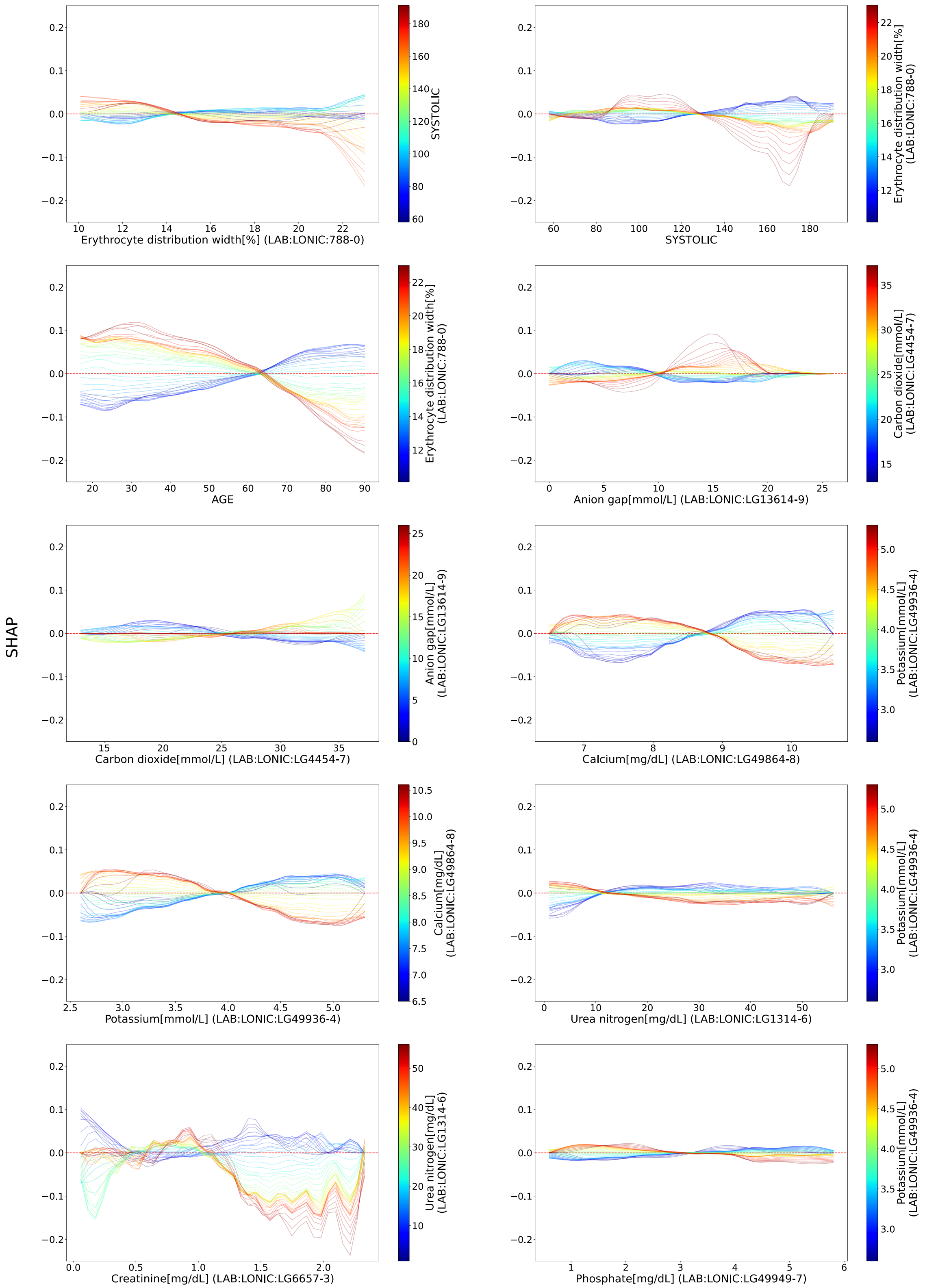


**Figure S9: Continuous-continuous bivariate analysis**. This figure illustrates only the interaction term of the bivariate analysis.

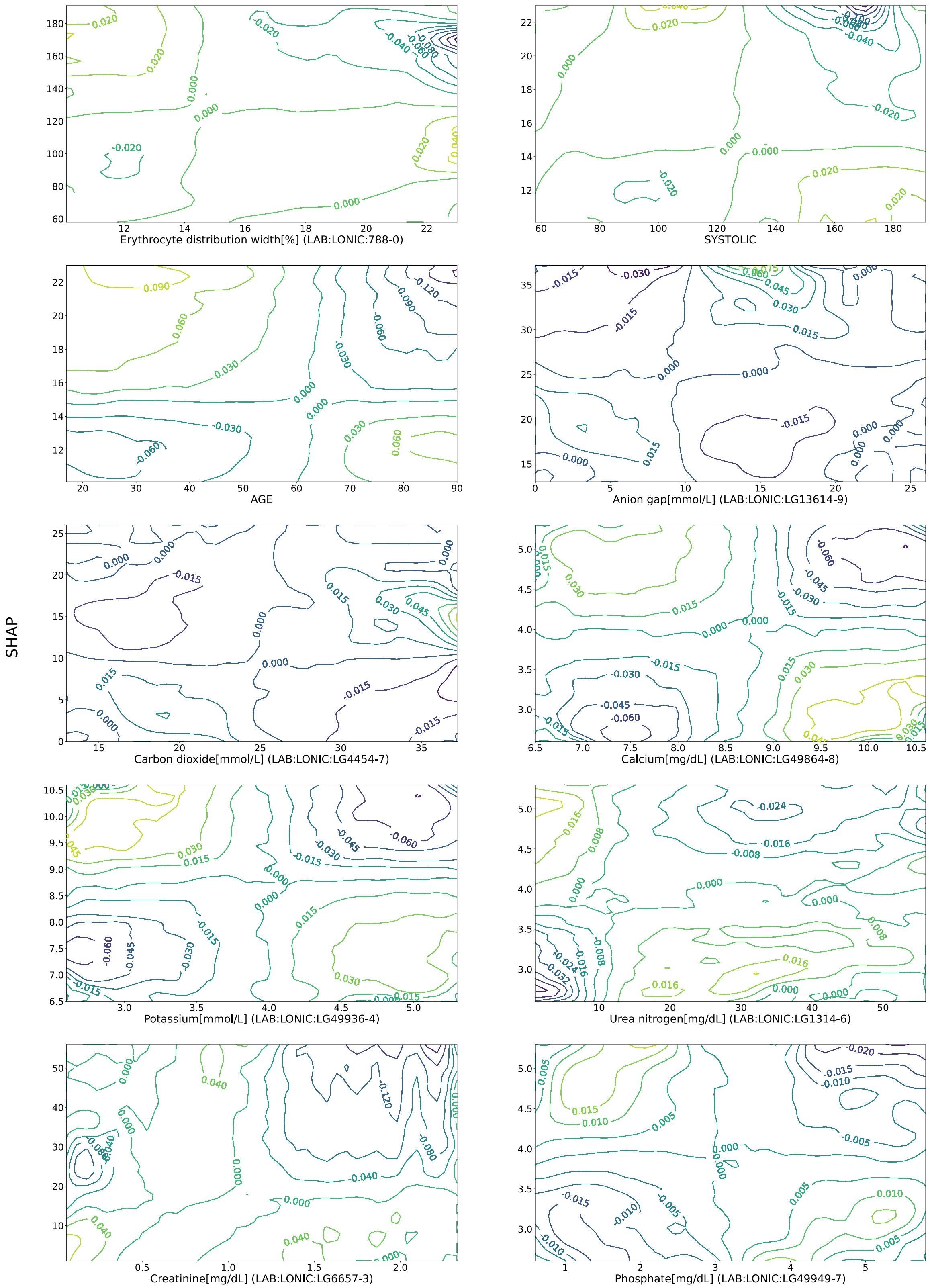


**Figure S10: Continuous-continuous bivariate contour maps**. This figure illustrates only the interaction term of the bivariate analysis in contour graph.

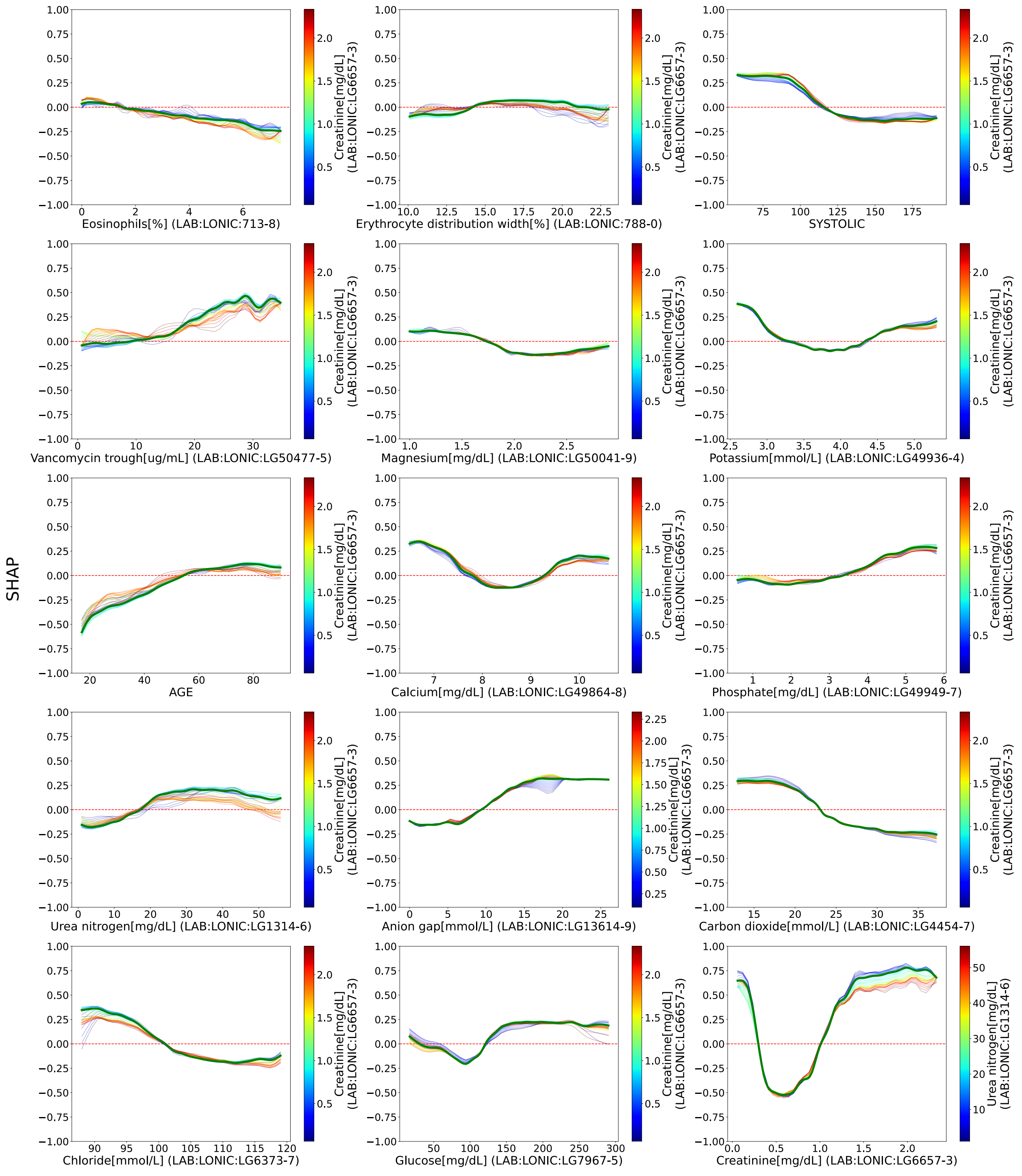


**Figure S11: Continuous-continuous bivariate analysis**. This figure illustrates the interaction between two continuous variables. It highlights pairs with the greatest improvement in $r^2$ compared to the univariate $r^2$ of the primary features.

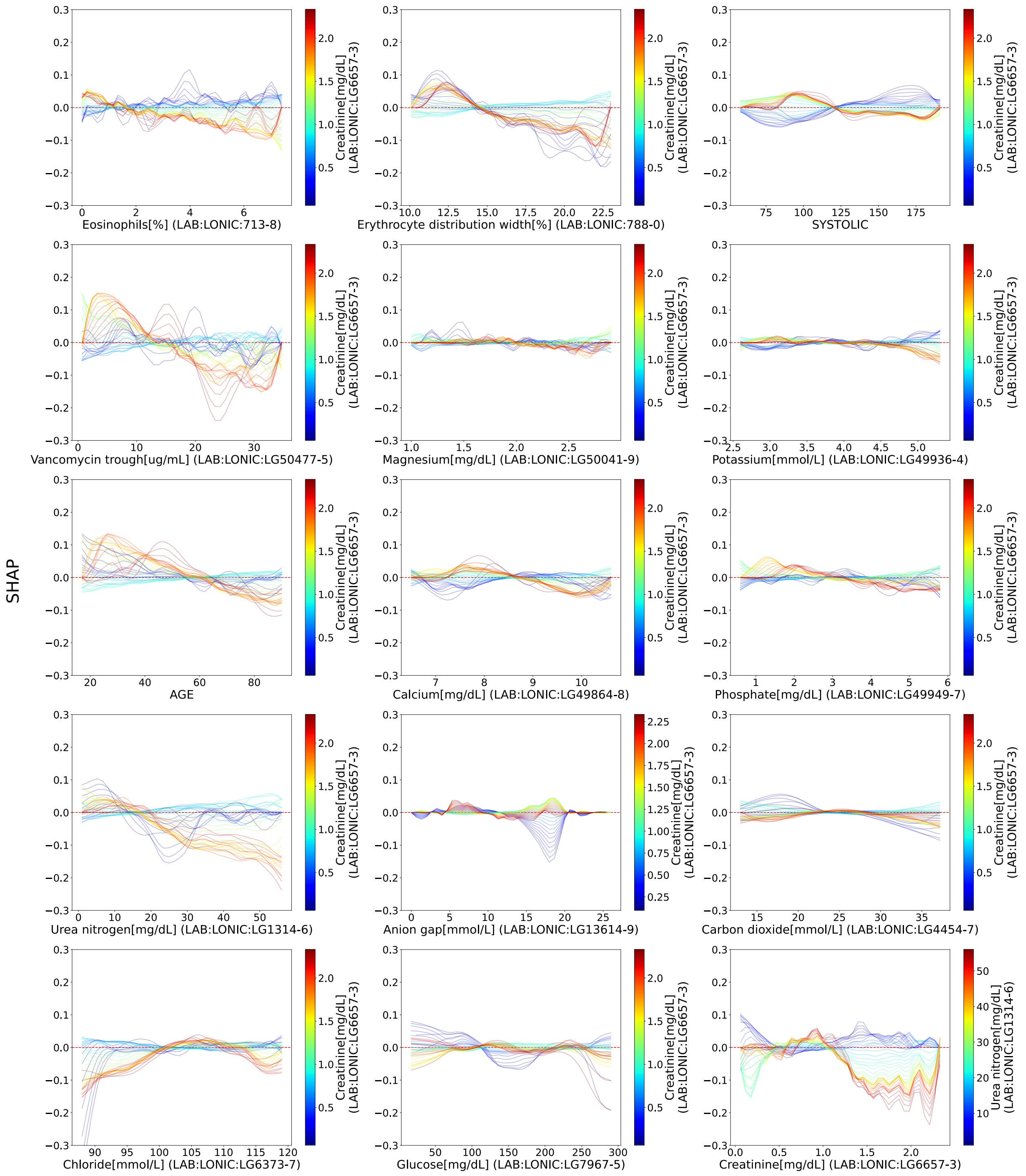


**Figure S12: Continuous-continuous bivariate analysis (cont.)**. This figure illustrates only the interaction term of the bivariate analysis in contour graph.

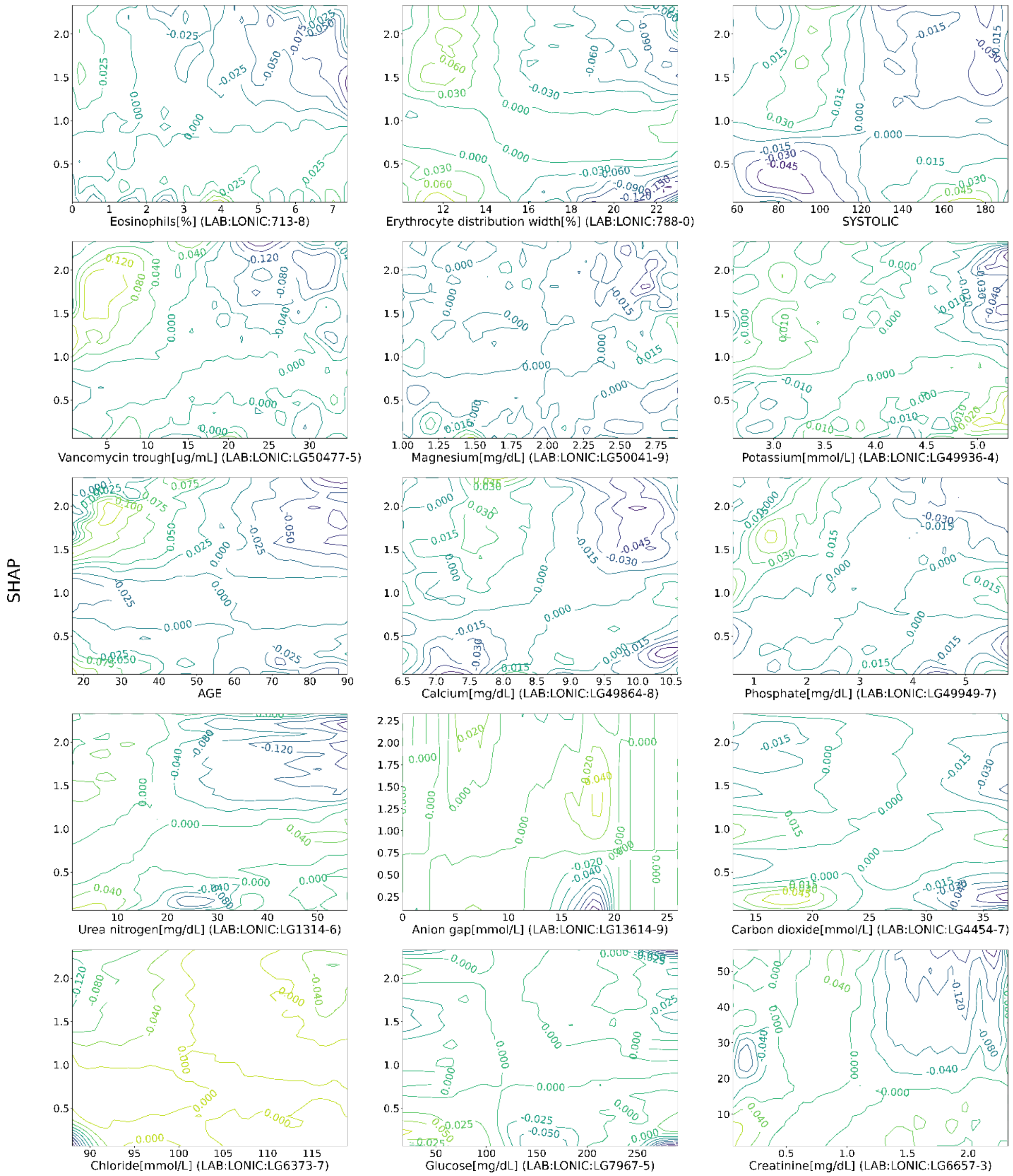


**Figure S13: Continuous-continuous bivariate contour maps**. This figure illustrates only the interaction term of the bivariate analysis.

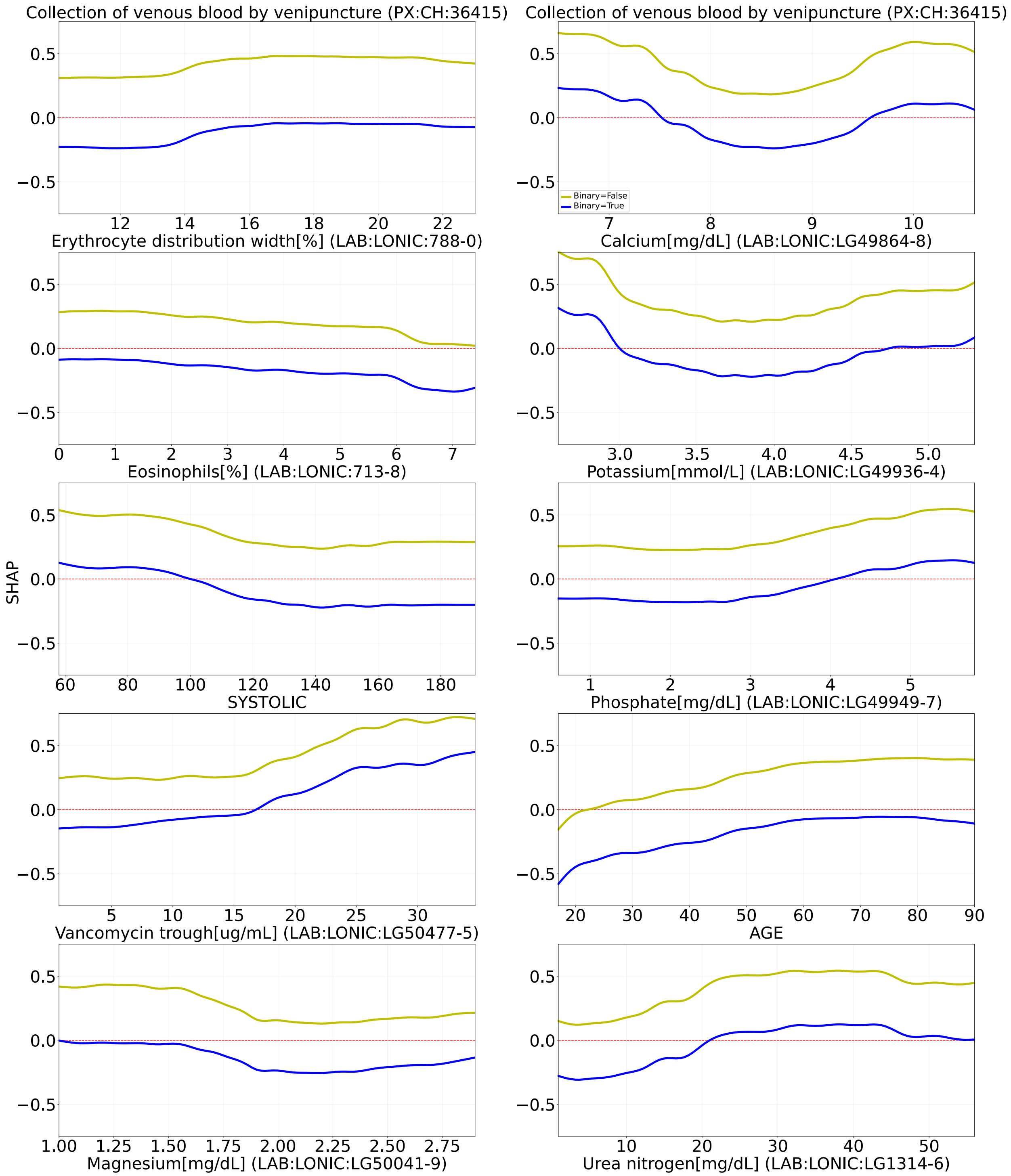


**Figure S14: Discrete-continuous bivariate analysis**. This figure illustrates the interaction between a continuous and a binary variable. It highlights pairs with the greatest improvement in $r^2$ compared to the maximum of univariate $r^2$ of the two predictors. Two curves in each panel correspond to cases where the binary variable is true (blue), false (yellow). All legends are the same and displayed on the top right. The continuous variable is indicated on the x-axis, while the discrete variable is shown as the column title.

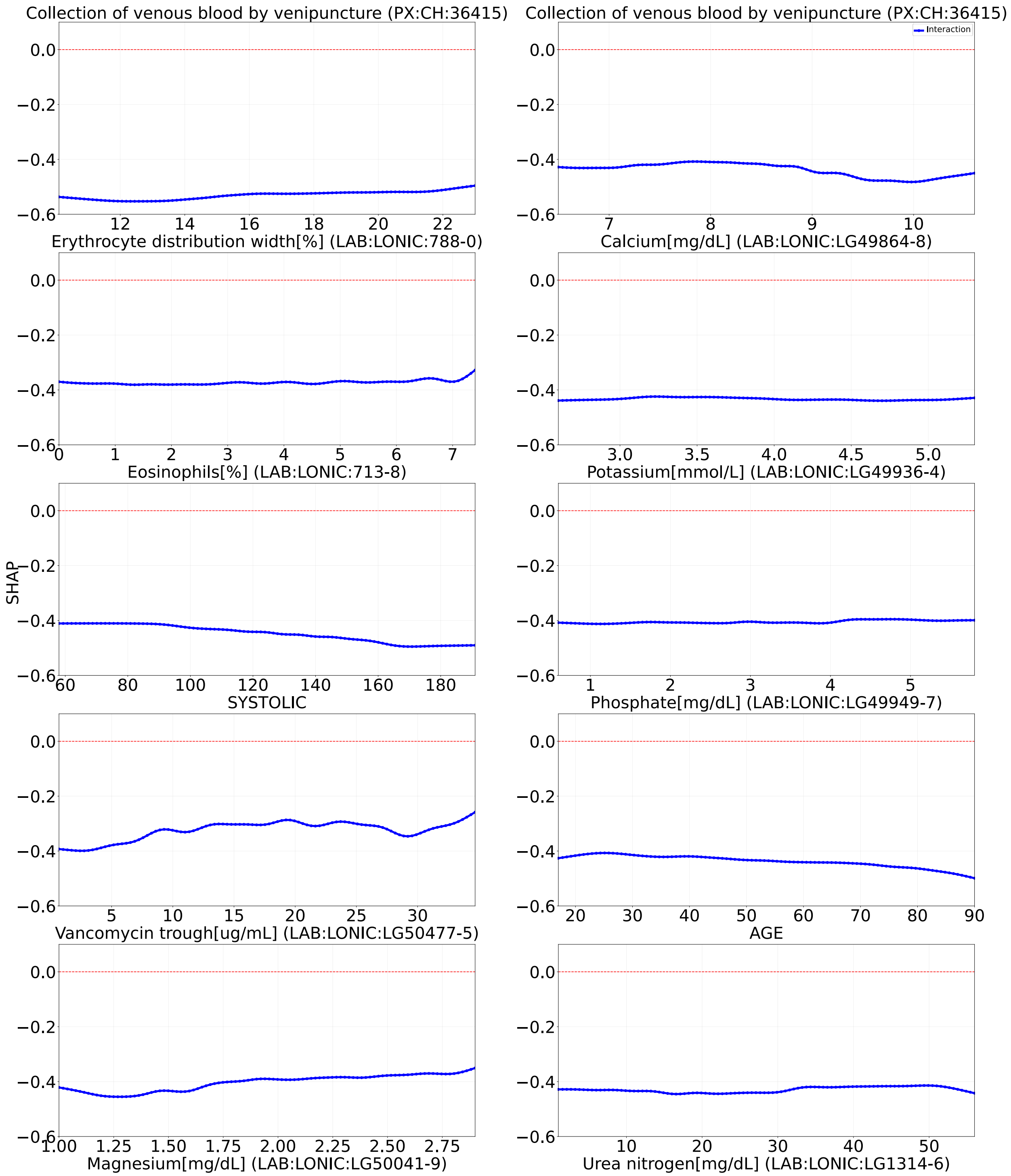


**Figure S15: Discrete-continuous bivariate analysis (cont.)**. This figure illustrates the difference between the positive and negative curve in **Figure S13**.

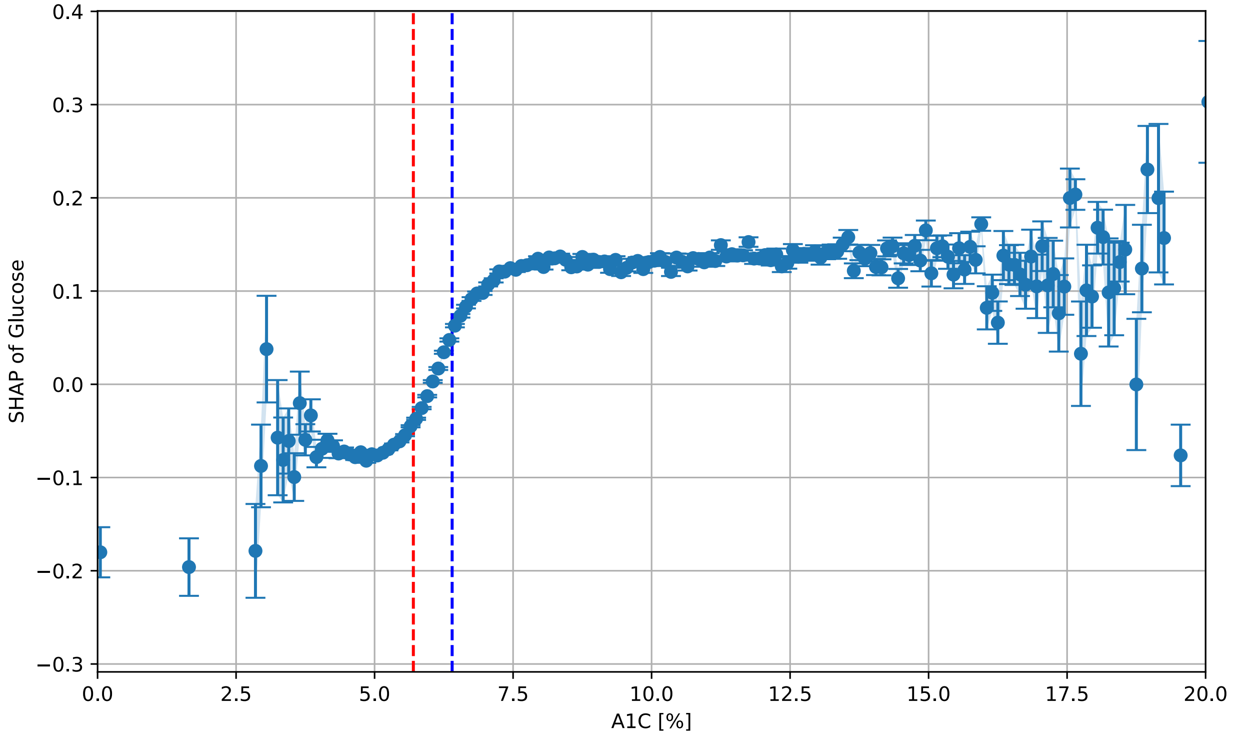


**Figure S16: SHAP value of glucose vs corresponding A1c**. This figure plots the glucose SHAP value of the individual encounter against the most recently known A1c. The vertical lines show the range of prediabetes A1c.

**Table S1: Details of the medical codes in this study.**

| Name | Code | Type |
|---|---|---|
| **Albumin** | LG61151-7 | Laboratory (LOINC) |
| **Anion gap** | LG13614-9 | Laboratory (LOINC) |
| **Anion gap 3** | 10466-1 | Laboratory (LOINC) |
| **Anion gap 4** | LG6139-2 | Laboratory (LOINC) |
| **Bicarbonate** | LG2807-8 | Laboratory (LOINC) |
| **Calcium** | LG49864-8 | Laboratory (LOINC) |
| **Carbon dioxide** | LG4454-7 | Laboratory (LOINC) |
| **Chloride** | LG6373-7 | Laboratory (LOINC) |
| **Creatinine** | LG50024-5 | Laboratory (LOINC) |
| **Deprecated bicarbonate** | 1962-0 | Laboratory (LOINC) |
| **Eosinophils** | LG32849-8 | Laboratory (LOINC) |
| **Erythrocyte distribution width** | LG788-0 | Laboratory (LOINC) |
| **Erythrocytes** | LG32850-6 | Laboratory (LOINC) |
| **Glucose** | LG7967-5 | Laboratory (LOINC) |
| **Granulocytes** | LG19023-1 | Laboratory (LOINC) |
| **Hematocrit** | LG4544-3 | Laboratory (LOINC) |
| **Hemoglobin** | LG44868-4 | Laboratory (LOINC) |
| **INR in platelet poor plasma** | LG6301-6 | Laboratory (LOINC) |
| **Leukocytes** | LG32857-1 | Laboratory (LOINC) |
| **Lymphocytes** | LG32863-9 | Laboratory (LOINC) |
| **Magnesium** | LG5903-2 | Laboratory (LOINC) |
| **MCHC** | LG786-4 | Laboratory (LOINC) |
| **MCV** | LG787-2 | Laboratory (LOINC) |
| **Monocytes** | LG5905-5 | Laboratory (LOINC) |
| **Neutrophils** | LG770-8 | Laboratory (LOINC) |
| **Osmolality** | 18182-6 | Laboratory (LOINC) |
| **Phosphate** | LG6426-3 | Laboratory (LOINC) |
| **Platelet mean volume** | LG32623-1 | Laboratory (LOINC) |
| **Platelets** | LG32892-8 | Laboratory (LOINC) |
| **Potassium** | LG49936-4 | Laboratory (LOINC) |
| **Segmented neutrophils** | LG26505-8 | Laboratory (LOINC) |
| **Sodium** | LG11363-5 | Laboratory (LOINC) |
| **Urea nitrogen** | LG1314-6 | Laboratory (LOINC) |
| **Vancomycin** | LG47183-5 | Laboratory (LOINC) |
| **Vancomycin trough** | LG50477-5 | Laboratory (LOINC) |
| **Aldosterone antagonists** | C03DA | Medication (ATC) |
| **Angiotensin II receptor blockers (ARBs)** | C09DX | Medication (ATC) |
| **Anilides** | N02BE | Medication (ATC) |
| **Antipropulsives** | A07DA | Medication (ATC) |
| **Bisphosphonates** | M05BA | Medication (ATC) |
| **Glucocorticoid** | H02AB | Medication (ATC) |
| **Glycopeptide antibacterials** | J01XA | Medication (ATC) |
| **Heparin group** | B01AB | Medication (ATC) |
| **Platelet aggregation inhibitors** | B01AC | Medication (ATC) |
| **Renin-inhibitor** | C09XA | Medication (ATC) |
| **Sulfonamides, plain** | C03CA | Medication (ATC) |
| **Thiazides, plain** | C03AA | Medication (ATC) |
| **Vasopressin antagonist** | C03XA | Medication (ATC) |
| **Collection of venous blood by venipuncture** | 36415 | Procedure (CPT) |
| **Dexamethasone sodium phosphate** | J1100 | Procedure (CPT) |
| **Diagnostic ultrasound procedures** | J3490 | Procedure (CPT) |
| **Injection, furosemide** | J1940 | Procedure (CPT) |
| **Injection, piperacillin sodium/tazobactam sodium** | J2543 | Procedure (CPT) |

| | | |
|---|---|---|
| **Self-care/home management training** | 97535 | Procedure (CPT) |
| **Transparent film, sterile** | A6257 | Procedure (CPT) |
| **Unclassified drugs** | J3490 | Procedure (CPT) |

**Table S2: Number of predictors used in final model per site.**

| Site | 1 | 2 | 3 | 4 | 5 | 6 | 7 | 8 | 9 |
|---|---|---|---|---|---|---|---|---|---|
| # of Features | 3011 | 1426 | 1016 | 689 | 1007 | 1264 | 1014 | 514 | 1642 |

**Table S3: Linear fitting results.**

| | Intercept | Slope | $R^2$ |
|---|---|---|---|
| Creatinine[mg/dL] (LAB:LONIC:LG6657-3) | -1.1276414 | 1.0846594 | 0.7339211 |
| Anion gap[mmol/L] (LAB:LONIC:LG13614-9) | -0.3789561 | 0.0419410 | 0.7035009 |
| Chloride[mmol/L] (LAB:LONIC:LG6373-7) | 2.7145285 | -0.0267053 | 0.6962211 |
| Glucose[mg/dL] (LAB:LONIC:LG7967-5) | -0.5000911 | 0.0038513 | 0.6510272 |
| Carbon dioxide[mmol/L] (LAB:LONIC:LG4454-7) | 0.8176410 | -0.0349743 | 0.6465330 |
| Urea nitrogen[mg/dL] (LAB:LONIC:LG1314-6) | -0.2284675 | 0.0128684 | 0.6361570 |
| Phosphate[mg/dL] (LAB:LONIC:LG49949-7) | -0.3277952 | 0.1061907 | 0.6099552 |
| AGE | -0.4840450 | 0.0079214 | 0.5628581 |
| Eosinophils[%] (LAB:LONIC:713-8) | 0.0516463 | -0.0364750 | 0.5140343 |
| Magnesium[mg/dL] (LAB:LONIC:LG50041-9) | 0.3600246 | -0.2197038 | 0.4758192 |
| Vancomycin trough[ug/mL] (LAB:LONIC:LG50477-5) | -0.1948305 | 0.0202784 | 0.4680749 |
| Monocytes[%] (LAB:LONIC:5905-5) | 0.0756002 | -0.0115843 | 0.4519267 |
| SYSTOLIC | 0.6614316 | -0.0054747 | 0.4158297 |
| BMI | -0.2001828 | 0.0061154 | 0.4099001 |
| MCHC[g/dL] (LAB:LONIC:786-4) | 0.5437077 | -0.0166329 | 0.3675092 |
| Lymphocytes[10*3/uL] (LAB:LONIC:LG32863-9) | -0.0192586 | -0.0000001 | 0.3544211 |
| Prothrombin time[s] (PT) | -0.0212156 | 0.0024368 | 0.3481438 |
| Erythrocyte distribution width[%] (LAB:LONIC:788-0) | -0.2733819 | 0.0184494 | 0.3069723 |
| Hematocrit[%] (LAB:LONIC:4544-3) | -0.1289703 | 0.0038169 | 0.2967586 |
| Leukocytes[10*3/uL] (LAB:LONIC:LG32857-1) | -0.0309337 | 0.0000000 | 0.2344047 |
| Bicarbonate[mmol/L] (LAB:LONIC:LG2807-8) | 0.1903741 | -0.0070700 | 0.2280318 |
| Neutrophils[10*3/uL] (LAB:LONIC:LG32886-0) | -0.0066886 | -0.0000000 | 0.2049124 |
| Potassium[mmol/L] (LAB:LONIC:LG49936-4) | -0.2939866 | 0.0661937 | 0.2026063 |
| Erythrocytes[10*6/uL] (LAB:LONIC:LG32850-6) | -0.0766517 | 0.0213843 | 0.1984652 |
| Hemoglobin[g/dL] (LAB:LONIC:LG44868-4) | -0.0779781 | 0.0073445 | 0.1518716 |
| Platelets[10*3/uL] (LAB:LONIC:LG32892-8) | -0.0178437 | -0.0000000 | 0.1485041 |
| DIASTOLIC | 0.0041803 | -0.0003852 | 0.1217182 |
| Sodium[mmol/L] (LAB:LONIC:LG11363-5) | -0.1079815 | 0.0007071 | 0.0954123 |
| Calcium[mg/dL] (LAB:LONIC:LG49864-8) | -0.3914282 | 0.0397104 | 0.0900969 |

**Table S4: Quadratic fitting results.**

| | Intercept | Slope | Curvature | $R^2$ |
|---|---|---|---|---|
| Chloride[mmol/L] (LAB:LONIC:LG6373-7) | 17.1667489 | -0.3073372 | 0.0013594 | 0.7729691 |
| Creatinine[mg/dL] (LAB:LONIC:LG6657-3) | -0.9042534 | 0.5950832 | 0.2340142 | 0.7424414 |
| Glucose[mg/dL] (LAB:LONIC:LG7967-5) | -0.9673367 | 0.0108372 | -0.0000238 | 0.7171592 |
| Carbon dioxide[mmol/L] (LAB:LONIC:LG4454-7) | 2.2862322 | -0.1516962 | 0.0022709 | 0.7169816 |
| Anion gap[mmol/L] (LAB:LONIC:LG13614-9) | -0.3298240 | 0.0310277 | 0.0005278 | 0.7047333 |
| Urea nitrogen[mg/dL] (LAB:LONIC:LG1314-6) | -0.3314335 | 0.0257195 | -0.0002998 | 0.6969313 |
| Phosphate[mg/dL] (LAB:LONIC:LG49949-7) | -0.0212457 | -0.0943111 | 0.0307429 | 0.6686879 |
| AGE | -0.8995648 | 0.0245017 | -0.0001484 | 0.6410513 |
| Calcium[mg/dL] (LAB:LONIC:LG49864-8) | 12.1914748 | -2.8845091 | 0.1690424 | 0.6028460 |
| Potassium[mmol/L] (LAB:LONIC:LG49936-4) | 3.5673968 | -1.8948053 | 0.2460578 | 0.6014966 |
| Magnesium[mg/dL] (LAB:LONIC:LG50041-9) | 1.4760552 | -1.3687754 | 0.2901568 | 0.5905891 |
| SYSTOLIC | 2.6115090 | -0.0362603 | 0.0001190 | 0.5189142 |
| Eosinophils[%] (LAB:LONIC:713-8) | 0.0468984 | -0.0287646 | -0.0014622 | 0.5169378 |
| Erythrocyte distribution width[%] (LAB:LONIC:788-0) | -1.7900698 | 0.2097699 | -0.0058926 | 0.5095903 |
| Vancomycin trough[ug/mL] (LAB:LONIC:LG50477-5) | -0.0764602 | 0.0032932 | 0.0005218 | 0.4818887 |
| Monocytes[%] (LAB:LONIC:5905-5) | 0.0679902 | -0.0094139 | -0.0001304 | 0.4527882 |
| Prothrombin time[s] (PT) | -0.3198388 | 0.0371128 | -0.0009030 | 0.4350250 |
| BMI | -0.0292774 | -0.0057365 | 0.0001943 | 0.4320248 |
| Lymphocytes[10*3/uL] (LAB:LONIC:LG32863-9) | -0.0160534 | -0.0000003 | 0.0000000 | 0.3969037 |
| DIASTOLIC | 1.7837614 | -0.0510090 | 0.0003519 | 0.3849066 |
| MCHC[g/dL] (LAB:LONIC:786-4) | 0.5340051 | -0.0160420 | -0.0000090 | 0.3675093 |
| Sodium[mmol/L] (LAB:LONIC:LG11363-5) | 30.5256190 | -0.4453097 | 0.0016224 | 0.3537723 |
| Hematocrit[%] (LAB:LONIC:4544-3) | -0.0951296 | 0.0017318 | 0.0000310 | 0.2973749 |
| Leukocytes[10*3/uL] (LAB:LONIC:LG32857-1) | -0.0359116 | 0.0000001 | -0.0000000 | 0.2606312 |
| Bicarbonate[mmol/L] (LAB:LONIC:LG2807-8) | 0.4412796 | -0.0269776 | 0.0003833 | 0.2357406 |
| Neutrophils[10*3/uL] (LAB:LONIC:LG32886-0) | -0.0060810 | -0.0000000 | 0.0000000 | 0.2093275 |
| Erythrocytes[10*6/uL] (LAB:LONIC:LG32850-6) | -0.0522640 | 0.0079270 | 0.0017859 | 0.1991024 |
| Hemoglobin[g/dL] (LAB:LONIC:LG44868-4) | -0.1503240 | 0.0207021 | -0.0005936 | 0.1565820 |
| Platelets[10*3/uL] (LAB:LONIC:LG32892-8) | 0.0000000 | -0.0000000 | 0.0000000 | 0.1549557 |

**Table S5: Spline fitting results.**

| | Intercept | $R^2$ |
|---|---|---|
| Creatinine[mg/dL] (LAB:LONIC:LG6657-3) | -0.1964600 | 0.9152868 |
| Glucose[mg/dL] (LAB:LONIC:LG7967-5) | -0.0284707 | 0.8478003 |
| Chloride[mmol/L] (LAB:LONIC:LG6373-7) | -0.0513699 | 0.7974916 |
| Carbon dioxide[mmol/L] (LAB:LONIC:LG4454-7) | -0.0722191 | 0.7500038 |
| Urea nitrogen[mg/dL] (LAB:LONIC:LG1314-6) | -0.0277501 | 0.7441468 |
| Anion gap[mmol/L] (LAB:LONIC:LG13614-9) | 0.0459761 | 0.7308167 |
| Phosphate[mg/dL] (LAB:LONIC:LG49949-7) | 0.0128489 | 0.6835476 |
| Potassium[mmol/L] (LAB:LONIC:LG49936-4) | -0.0336439 | 0.6676370 |
| Calcium[mg/dL] (LAB:LONIC:LG49864-8) | -0.0478868 | 0.6650750 |
| AGE | -0.0161394 | 0.6494442 |
| Magnesium[mg/dL] (LAB:LONIC:LG50041-9) | -0.0723084 | 0.6435627 |
| Erythrocyte distribution width[%] (LAB:LONIC:788-0) | 0.0061504 | 0.5449173 |
| SYSTOLIC | -0.0293939 | 0.5313162 |
| Eosinophils[%] (LAB:LONIC:713-8) | -0.0145281 | 0.5299112 |
| Vancomycin trough[ug/mL] (LAB:LONIC:LG50477-5) | 0.1106271 | 0.5058633 |
| Monocytes[%] (LAB:LONIC:5905-5) | -0.0184799 | 0.4811464 |
| Prothrombin time[s] (PT) | 0.0170509 | 0.4655078 |
| BMI | -0.0248238 | 0.4588218 |
| DIASTOLIC | -0.0258824 | 0.4260924 |
| Lymphocytes[10*3/uL] (LAB:LONIC:LG32863-9) | -0.0323234 | 0.4246160 |
| Sodium[mmol/L] (LAB:LONIC:LG11363-5) | -0.0105833 | 0.3961822 |
| MCHC[g/dL] (LAB:LONIC:786-4) | -0.0073202 | 0.3948949 |
| Hematocrit[%] (LAB:LONIC:4544-3) | -0.0022561 | 0.3248959 |
| Leukocytes[10*3/uL] (LAB:LONIC:LG32857-1) | -0.0147896 | 0.2733390 |
| Bicarbonate[mmol/L] (LAB:LONIC:LG2807-8) | 0.0082736 | 0.2495821 |
| Neutrophils[10*3/uL] (LAB:LONIC:LG32886-0) | -0.0106107 | 0.2132900 |
| Erythrocytes[10*6/uL] (LAB:LONIC:LG32850-6) | 0.0030121 | 0.2087695 |
| Hemoglobin[g/dL] (LAB:LONIC:LG44868-4) | 0.0025512 | 0.1678970 |
| Platelets[10*3/uL] (LAB:LONIC:LG32892-8) | -0.0208653 | 0.1587638 |

**Table S6: Results from confounding analysis.**

| | **Low Potassium** | **High Potassium** | **Low Sodium** | **High Calcium** |
|---|---|---|---|---|
| **$\beta$ from Eq. (1)** | 0.085* | 0.0274* | -0.0002 | 0.0332* |
| **$\beta'$ from Eq. (2)** | 0.0352* | 0.0778* | -0.0007* | 0.0329* |
| **change** | 0.5859 | 1.8371 | 3.1922 | 0.8754 |

Note: * represents $p < 0.05$.

**Table S7: Fitting characteristics (Interaction).**

| Primary | Secondary | Multi $R^2$ | Primary $R^2$ | Secondary $R^2$ |
|---|---|---|---|---|
| **Creatinine[mg/dL]** | Urea nitrogen[mg/dL] | 0.9209162 | 0.9152868 | 0.7441468 |
| **Anion gap[mmol/L]** | Carbon dioxide[mmol/L] | 0.7717843 | 0.7308167 | 0.7500038 |
| **Carbon dioxide[mmol/L]** | Anion gap[mmol/L] | 0.7717843 | 0.7500038 | 0.7308167 |
| **Urea nitrogen[mg/dL]** | Potassium[mmol/L] | 0.7515306 | 0.7441468 | 0.6676370 |
| **Age** | Erythrocyte distribution width[%] | 0.7095151 | 0.6494442 | 0.5449173 |
| **Phosphate[mg/dL]** | Potassium[mmol/L] | 0.6856414 | 0.6835476 | 0.6676370 |
| **Calcium[mg/dL]** | Potassium[mmol/L] | 0.6769515 | 0.6650750 | 0.6676370 |
| **Potassium[mmol/L]** | Calcium[mg/dL] | 0.6769515 | 0.6676370 | 0.6650750 |
| **Erythrocyte distribution width[%]** | Systolic blood pressure | 0.6223338 | 0.5449173 | 0.5313162 |
| **Systolic blood pressure** | Erythrocyte distribution width[%] | 0.6223338 | 0.5313162 | 0.5449173 |

**Table S8: Fitting characteristics (Interaction) [Collection of venous blood by venipuncture].**

| | Intercept | Interaction Intercept | $R^2$ |
|---|---|---|---|
| **LAB:LG6657-3(mg/dL)** | 0.0976566 | -0.4089278 | 0.8771373 |
| **LAB:788-0(%)** | 0.4072924 | -0.5384763 | 0.8572973 |
| **LAB:LG49949-7(mg/dL)** | 0.3048430 | -0.4063698 | 0.7854123 |
| **LAB:LG7967-5(mg/dL)** | 0.2852514 | -0.4374054 | 0.7832703 |
| **LAB:LG1314-6(mg/dL)** | 0.2867546 | -0.4357481 | 0.7827167 |
| **LAB:LG49864-8(mg/dL)** | 0.2699381 | -0.4308862 | 0.7796226 |
| **DIASTOLIC** | 0.2976216 | -0.4471331 | 0.7789009 |
| **LAB:LG49936-4(mmol/L)** | 0.2694806 | -0.4316616 | 0.7770885 |
| **LAB:LG50041-9(mg/dL)** | 0.2069354 | -0.3976390 | 0.7730089 |
| **LAB:LG6373-7(mmol/L)** | 0.2531592 | -0.4316577 | 0.7549775 |
| **ORIGINAL_BMI** | 0.3005586 | -0.4467059 | 0.7519202 |
| **AGE** | 0.3055611 | -0.4422391 | 0.7512631 |
| **LAB:LG4454-7(mmol/L)** | 0.1669962 | -0.3270793 | 0.7475802 |
| **LAB:713-8(%)** | 0.2549172 | -0.3747820 | 0.7451524 |
| **SYSTOLIC** | 0.2993576 | -0.4466522 | 0.7261778 |
| **LAB:LG13614-9(mmol/L)** | 0.2935247 | -0.3923937 | 0.6739725 |
| **LAB:LG50477-5(ug/mL)** | 0.3329021 | -0.3156269 | 0.6736715 |
